# A Randomized, Double-Blind, Placebo-Controlled, Single Ascending Oral Dose Study of Mocravimod: Safety, Tolerability, Pharmacokinetics, and Pharmacodynamics in Healthy Participants

**DOI:** 10.64898/2026.05.11.26352861

**Authors:** Dymphy Huntjens, Dirk Klingbiel, Jens Hasskarl

**Affiliations:** Priothera SAS, Saint-Louis, France

**Keywords:** Mocravimod, immunomodulator, pharmacokinetics, single ascending dose, healthy volunteers

## Abstract

Mocravimod (KRP203) is a selective sphingosine 1-phosphate (S1P) receptor modulator currently in development for patients with haematological malignancies undergoing allogenic haematopoietic cell transplantation (HCT). This first-in-human, randomised, double-blind, placebo-controlled, single ascending oral dose study evaluated the safety, tolerability, pharmacokinetics (PK), and pharmacodynamics (PD) of mocravimod in 136 healthy adult participants (EudraCT No. 2006-006814-13).

Participants received single doses ranging from 0.01 to 40 mg or placebo, with a cohort dedicated to studying food-effect at 3 mg. Mocravimod demonstrated slow absorption (mean T_max_ 6–11 hrs), extensive distribution, and a long terminal half-life (91–132 hrs). Exposure increased dose-proportionally for doses ≥2 mg. The most common adverse events were headache, dizziness, and fatigue, all graded as mild or moderate; no serious adverse events or deaths occurred. Mocravimod-phosphate induced robust, dose-dependent reductions in lymphocyte counts, with significant decreases at doses ≥2 mg and recovery to baseline observed in all but the highest dose groups. Cardiac effects included transient bradycardia and benign second-degree atrioventricular (AV) block at higher doses, without clinically significant arrhythmias. Food intake had minimal impact on PK. No clinically meaningful changes in pulmonary function or laboratory safety signals were detected.

These results indicate that single oral doses of mocravimod up to 40 mg are safe and well tolerated in healthy adults, with predictable PK and expected PD effects. The findings support further clinical development of mocravimod as a targeted immunomodulator in settings such as allogeneic HCT for haematological malignancies.

## Introduction

Sphingosine-1-phosphate (S1P) receptor modulators have transformed the management of immune-mediated diseases such as multiple sclerosis (MS) by targeting lymphocyte trafficking. The first-in-class agent, fingolimod, demonstrated that functional antagonism of S1P isoform 1 (S1P1) receptor sequesters lymphocytes in lymphoid tissues, reducing their egress and thereby modulating immune responses. This mechanism underpins the efficacy of S1P modulators in MS and has led to the approval of several next-generation agents—siponimod, ozanimod, ponesimod, and etrasimod—for MS and ulcerative colitis, each with improved receptor selectivity and safety profiles [1–3]. Recent reviews highlight that S1P modulators are now being explored in a range of indications, including neurodegenerative and inflammatory diseases, due to their ability to modulate immune cell migration with very limited immunosuppression [1, 4].

Mocravimod (KRP203) is an S1P modulator with strong affinity towards S1P1/S1P5 [5]. Mocravimod is metabolized *in vivo* into its phosphorylated active metabolite mocravimod-phosphate. Unlike conventional immunosuppressants, S1P modulators do not impair T-cell function, including cytotoxicity, preserving anti-tumour immunity [6]. The potential mode of action is by sequestering lymphocytes to lymphatic tissue and bone marrow from the circulation. The prevention of lymphocyte egress has also been demonstrated for the bone marrow compartment in preclinical models [7]. Its mechanism of action has shown promise in preclinical models of autoimmunity, transplantation, and haematological malignancies [7].

Mocravimod is currently under development as an adjunctive and maintenance treatment for patients with acute myeloid leukaemia (AML) undergoing allogeneic haematopoietic cell transplantation (allo-HCT) (NCT05429632). Allo-HCT remains the only curative option for patients with intermediate or high-risk AML, leveraging the graft-versus-leukaemia (GvL) effect—where donor T cells eradicate residual malignant cells [8]. However, this benefit is counterbalanced by graft-versus-host disease (GvHD), a major cause of morbidity and mortality post-transplant, resulting from donor T-cell-mediated attack on host tissues [9, 10].

S1P modulators restrict T-cell egress from lymphoid tissues and offer a unique approach: they can limit the migration of alloreactive T cells to peripheral organs—mitigating the risk of GvHD—while preserving their anti-leukaemic activity within lymphoid and bone marrow compartments [11, 12]. Preclinical and early clinical data suggest that mocravimod may achieve this “decoupling” of GvHD and GvL, supporting its development as an adjunct to allo-HCT in AML [7, 13]. S1P receptor modulators such as mocravimod or fingolimod have shown beneficial therapeutic effects in preclinical models of GvHD [6, 14].

Based on the results from nonclinical studies, a first-in-human, Phase 1 study was designed to evaluate the safety, tolerability, and single-dose pharmacokinetics of mocravimod in healthy volunteers. The findings from this trial served as a basis for further clinical development of mocravimod in haematological malignancies.

## Materials and Methods

### Study Design

This single-centre, randomised, double-blind, placebo-controlled, parallel-group, ascending single oral dose study of mocravimod, was conducted in healthy adult volunteers. The study was designed in two parts: (1) single ascending dose escalation and (2) food effect assessment.

Part 1 was a randomized, double-blind, placebo-controlled, time-lagged, parallel-group design to evaluate the safety, tolerability, PK, and PD of single ascending doses (SADs) of mocravimod. Thirteen ascending dose levels (0.01, 0.03, 0.1, 0.3, 1, 2, 3, 4.5, 6, 9, 15, 25, and 40 mg) were assessed, with 10 participants (8 active, 2 placebo) randomly assigned to each cohort, except Cohort IX (13 active, 3 placebo). Actual dose escalation was based on safety, tolerability, PK, and PD data from preceding cohorts. Dosing in Cohort I was staggered for safety, with initial dosing of two participants (one active, one placebo), followed by two subgroups of four participants each, contingent on safety review (see Figure 1A).

**Figure 1:**
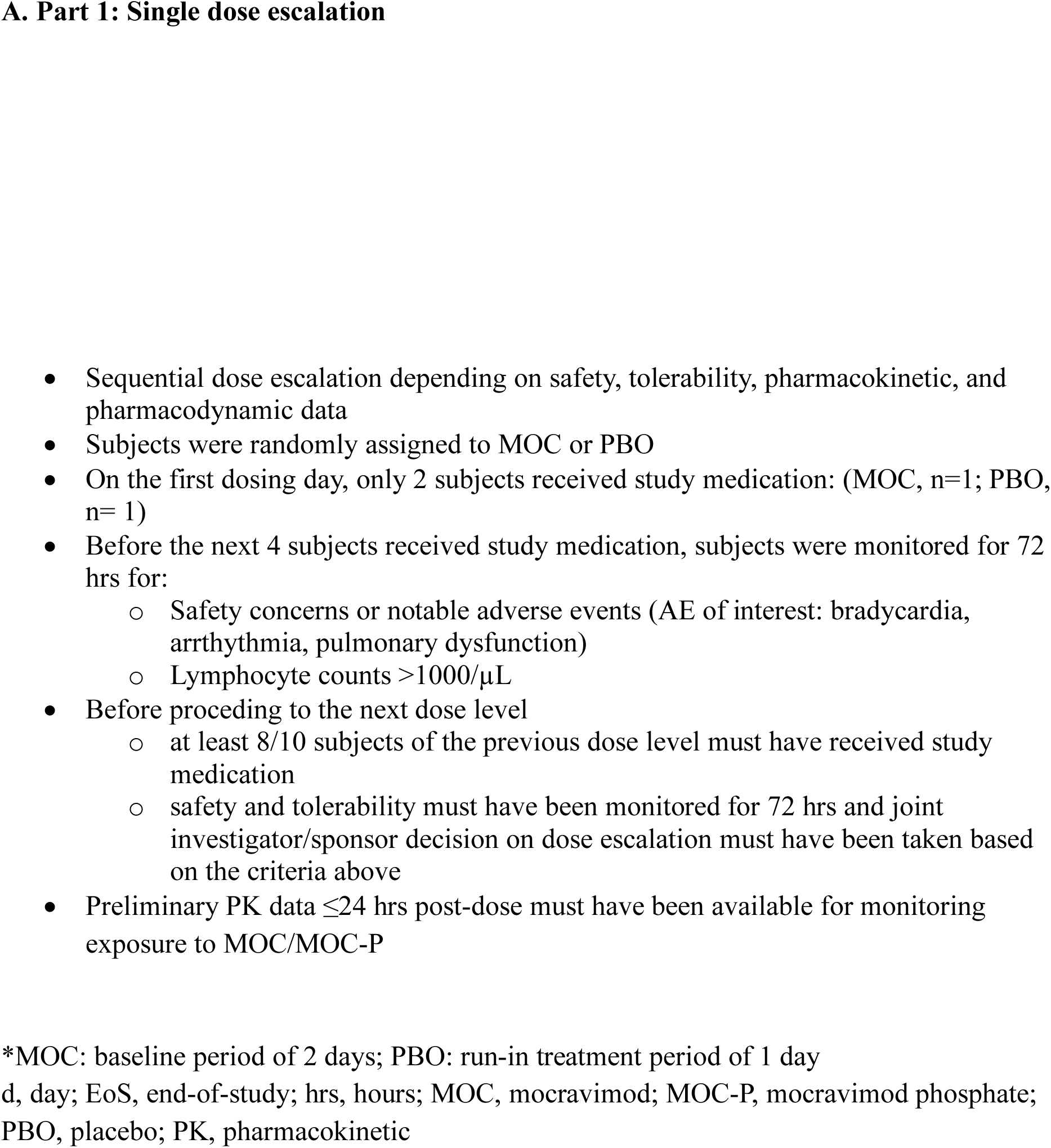
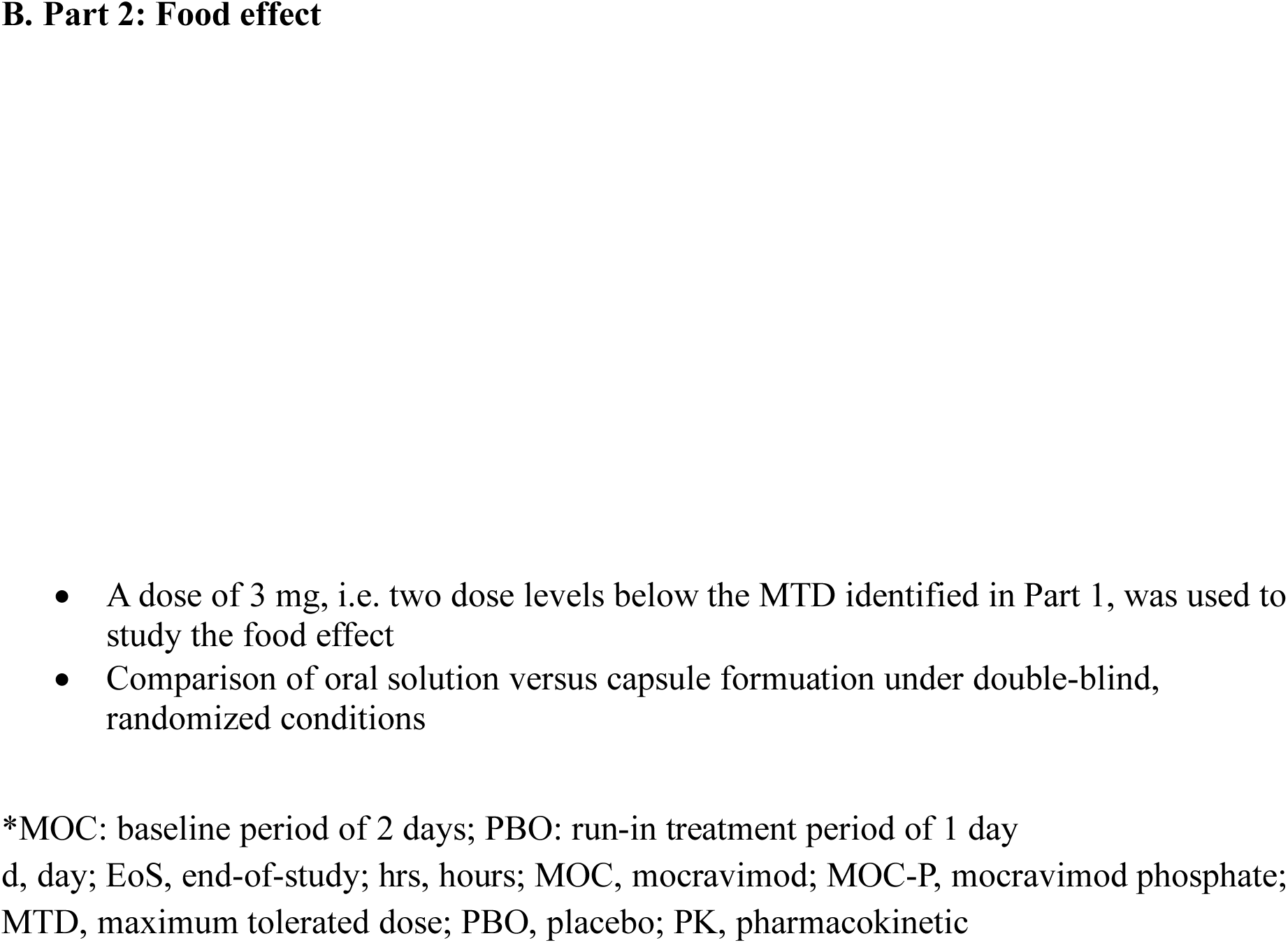
Study design.

Each participant entered a 20-day screening period (Day -22 to Day -3), followed by a baseline period (Day -2), a placebo run-in treatment period (Day -1), one single dose treatment with an in-house observation period of at least 96 hrs, two ambulatory visits (Day 7 and 10) and a study completion visit 14 days (±1 day) after drug administration (see Figure 1A).

Mocravimod was supplied as oral solution and as hard gelatine capsules (0.1 mg, 1 mg, 10 mg). Placebo capsules matched the active drug in appearance. Fasting was required for at least 10 hrs prior to dosing and for at least four hrs post-dose. Fluid intake was restricted around dosing, and meals were standardized during confinement.

Part 2 was an open-label, randomized, two-period, two-sequence crossover design to investigate the effect of food on the PK of a single 3 mg dose of mocravimod. The 3 mg capsule was administered under both fasted and fed conditions, with the fed condition involving a standard FDA breakfast 30 minutes prior to dosing with a washout period of at least three weeks between treatments (see Figure 1B).

Each participant entered a 20-day screening period (Day -21 to Day -2), followed by two baseline periods (Day -1), two single-dose treatment periods each with an in-house observation period of at least 96 hrs (study periods 1 and 2), three ambulatory visits in period 1 (Day 7, 10 and 15), two ambulatory visits in period 2 (Day 7 and 10) and a study completion visit 14 days (±1 day) after the last drug administration in period 2. The two single-dose treatments were separated by a wash-out period of at least 3 weeks (see Figure 1B).

### Study Population

Eligible participants were healthy male and female adults aged 18–55 years (inclusive), with a body mass index (BMI) of 18–30 kg/m² and a minimum weight of 60 kg. Participants were required to be in good health as determined by medical history, physical and neurological examination, vital signs, 12-lead electrocardiogram (ECG), and laboratory tests at screening. Female participants were required to be postmenopausal, surgically sterilized, or hysterectomized. Male participants were required to use double-barrier contraception and refrain from fathering a child for three months post-study. The main exclusion criteria were the use of tobacco products within three months prior to screening, recent participation in other clinical investigations or blood donation, a history or presence of cardiovascular, pulmonary, hepatic, renal, metabolic, or immunodeficiency diseases, forced expired volume in one second (FEV_1_) value below 85%, abnormal laboratory values, including white blood cell count outside 4,000–11,000/μL or platelets <100,000/μL.

### Blood Sampling and Bioanalysis

Blood samples (2 mL) for PK analysis were collected from a peripheral vein into potassium ethylenediaminetetraacetic acid (EDTA) tubes at pre-specified time points: pre-dose, 0.25, 0.5, 1, 2, 3, 4, 6, 8, 10, 12, 16, 24, 36, 48, 72, 96, 144, 216, and 336 hrs post-dose. Mocravimod and its active metabolite mocravimod-phosphate were quantified in whole blood using validated liquid chromatography-tandem mass spectrometry (LC-MS/MS) methods. The lower limit of quantification (LLOQ) was optimized during the study (0.5, 0.1, and 0.05 ng/mL for mocravimod; 0.5 and 0.1 ng/mL for mocravimod-phosphate). Not all dose levels were analysed with the same method due to ongoing optimization.

### Pharmacokinetic Parameters

Non-compartmental analysis was used to determine PK parameters such as area under the curve (AUC), AUC from zero to last measurable concentration (AUC_last_), AUC from zero to infinity (AUC_inf_), maximum concentration (C_max_), time to maximum concentration (T_max_), lag time (T_lag_), terminal elimination half-life (T½) and other PK parameters. Dose proportionality was assessed using a power model (log-transformed linear regression). Food effect was evaluated using a mixed-effects model adjusted for treatment, sequence, and period, with participant as a random effect.

### Pharmacodynamic and Safety Assessments

#### Lymphocyte Counts

Absolute lymphocyte counts were measured by a flow cytometer at pre-defined time points (Day -1 (period 1 only) and Day 1: pre-dose, 1, 2, 3, 4, 6, 8, 10, and 12 hrs post-dose, in the morning of Days 2, 3, 4 and 5 (i.e. 24, 48, 72, and 96 hrs after dosing on Day 1), in the morning of Days 7 and 10, Day 15 or study completion. PD parameters included nadir (maximum effect, E_max_), AUC, and time to nadir (TE_max_). Recovery to baseline and to 1000/µL was monitored until values returned to normal or study completion.

#### Cardiac Monitoring and Pulmonary Function

Heart rate was assessed by telemetry and 24-hr Holter ECG. Standard 12-lead ECGs were performed at specified intervals. Parameters included PR interval, QRS duration, QT interval (Bazett and Fridericia corrections), and rhythm analysis. Spirometry was performed according to American Thoracic Society (ATS)/European Respiratory Society (ERS) guidelines [15], measuring FEV_1_ and forced expiratory flow between 25% and 75% of exhaled vital capacity (FEF_25–75%_). Baseline and post-dose values (screening; both study periods: Day -1, Day 1: pre-dose, 1, 3, 6 and 12 hrs post-dose, on Days 5, 7 and 10, study completion) were compared, and changes from baseline were evaluated for clinical significance.

#### Supine Heart Rate Measurements

Supine heart rate was measured at screening, baseline (Day -2), Day -1, and on Day 1 at pre-dose, 0.5, 1, 1.5, 2, 3, 4, 6, 8, 10, 12, and 16 hrs post-dose, as well as in the mornings of Days 2 to 5 (i.e., 24, 48, 72, and 96 hrs after dosing on Day 1), and at the study completion visit.

Safety assessments included monitoring of AEs, clinical laboratory evaluations, serology, physical examination neurological examination, 12-lead ECG, vital signs (blood pressure, heart rate, respiratory rate, temperature), spirometry, breathlessness scores and exploratory special laboratory evaluations at prespecified intervals throughout the study.

### Ethics

The study was conducted at the PAREXEL Early Phase Clinical Unit (EPCU), London, United Kingdom (UK), in accordance with the Declaration of Helsinki, International Conference on Harmonisation Good Clinical Practice (ICH-GCP) guidelines, and all applicable regulatory requirements. The protocol, all amendments, and informed consent forms were reviewed and approved by the Brent Medical Ethics Committee, Northwick Park Hospital Harrow, Middlesex, UK, prior to study initiation. All participants provided written informed consent prior to any study-specific procedures (EudraCT No. 2006-006814-13).

### Statistical Methods

Descriptive statistics were used for all safety, PK, and PD endpoints. Dose proportionality was assessed using a log-transformed regression model. The food effect was analysed using a mixed-effects model. The minimum fraction from pre-dose (Emax) in absolute lymphocyte counts or heart rate was defined as the lowest post-Day 1, pre-dose value observed at any time point. Emax was summarized by treatment and analysed on the log scale using a linear effects model adjusted for log-transformed pre-dose values and mocravimod dose group. AUC under the lymphocyte counts during pre and post treatment period (over 0-24 hrs on Day -1 or Day 1) was calculated using the trapezoidal rule and analysed on the log scale. Geometric mean Emax and AUC values and geometric mean ratios versus placebo with 95% confidence intervals (CI) were estimated.

All participants who received at least one dose of study medication and had post-baseline data were included in the safety and PK/PD analysis sets.

## Results

### Baseline and Demographic Characteristics

A total of 136 healthy adult volunteers were enrolled in the study, with 135 completing Part 1 (single ascending dose) and 8 completing both periods of Part 2 (food effect). The study population in Part 1was predominantly male (99.3%), with one female participant. The median age was 29 years (range: 18–52), and the median weight was 77.2 kg (range: 60.5–102.0 kg) (Table 1). The majority of participants were Caucasian (64.7%), with representation from Black (13.2%), Asian (14%), Native American (1.5%), Pacific Islander (1.5%), and mixed ethnicities (5.1%). Baseline characteristics were well balanced across dose groups (data not shown). For Part 2, demographics were very similar to Part 1, e.g. the median age was 35 years (range: 21–51), and the median weight was 77.1 kg (range: 69.9–91.0) (Table 1).

**Table 1:** Patient demographics.

|  |  | **Part 1**<br>**SAD**<br>**N=136** | **Part 2**<br>**Food effect**<br>**N=9** |
| --- | --- | --- | --- |
| Age (years) | Mean (SD) | 30.9 (8.79) | 34.0 (10.14) |
|  | Median | 29.0 | 35.0 |
|  | Range | 18-52 | 21-51 |
| Sex – n (%) | Male | 135 (99.3%) | 9 (100%) |
|  | Female | 1 (0.7%) | 0 (0%) |
| Race – n (%) | Caucasian | 88 (64.7%) | 5 (55.6%) |
|  | Black | 18 (13.2%) | 4 (44.4%) |
|  | Asian | 19 (14%) | 0 (0%) |
|  | Native American | 2 (1.5%) | 0 (0%) |
|  | Pacific Islander | 2 (1.5%) | 0 (0%) |
|  | Other | 7 (5.1%) | 0 (0%) |
| Ethnicity – n (%) | Other | 106 (77.9%) | 9 (100%) |
|  | Hispanic/Latino | 3 (2.2%) | 0 (0%) |
|  | Chinese | 3 (2.2%) | 0 (0%) |
|  | Indian (Indian Subcontinent) | 12 (8.8%) | 0 (0%) |
|  | Mixed Ethnicity | 12 (8.8%) | 0 (0%) |
| Weight (kg) | Mean (SD) | 77.77 (9.613) | 80.11 (7.290) |
|  | Median | 77.20 | 77.10 |
|  | Range | 60.5-102.0 | 69.9-91.0 |
| Height (cm) | Mean (SD) | 178.1 (7.52) | 181.1 (4.73) |
|  | Median | 179.0 | 181.0 |
|  | Range | 154-199 | 175-192 |

SAD: single ascending dose; SD: standard deviation

### Pharmacokinetic SAD Results

Quantifiable blood concentrations for mocravimod and mocravimod-phosphate were observed for doses ≥2 mg. After drug intake, mocravimod was quantifiable after a lag-time of about 15 minutes to 1 hr, whereas mocravimod-phosphate showed a longer lag time ranging from about 30 minutes to 4 hrs. Concentrations of mocravimod peaked at about 6–11 hrs after drug intake and those of mocravimod phosphate peaked at 6–14 hrs. Blood concentrations of mocravimod-phosphate (median T_max_: 12 hrs) peaked later than those of mocravimod (median T_max_: 6 hrs). Both compounds showed a very slow elimination with a mono-exponential pattern. Blood concentrations appeared to increase proportionally to the dose (Figure 2). The exposure parameters from Part 1, displayed in Table 2A, C_max_, AUC_last_, and AUC_inf_ showed low to moderate variability (coefficient of variation [CV%] from 15% to less than 40%) in most groups. T_1/2_ was long and averaged 91–132 hrs. Like AUC_inf_, apparent clearance (CL/F) and volume of distribution (V/F) were better characterized for doses 6–40 mg. Distribution was extensive as suggested by V/F values of about 3000–6000 L. CL/F values fluctuated between 16.5 L/hr (minimal value in the 3 mg group) and 38.1 L/hr (maximal value in the 4.5 mg group). The geometric mean C_max_ ranged from 0.47 ng/mL (2 mg) to 15.9 ng/mL (40 mg), and AUC_inf_ from 32.9 hr*ng/mL (2 mg) to 1390 hr*ng/mL (40 mg).

**Figure 2:**
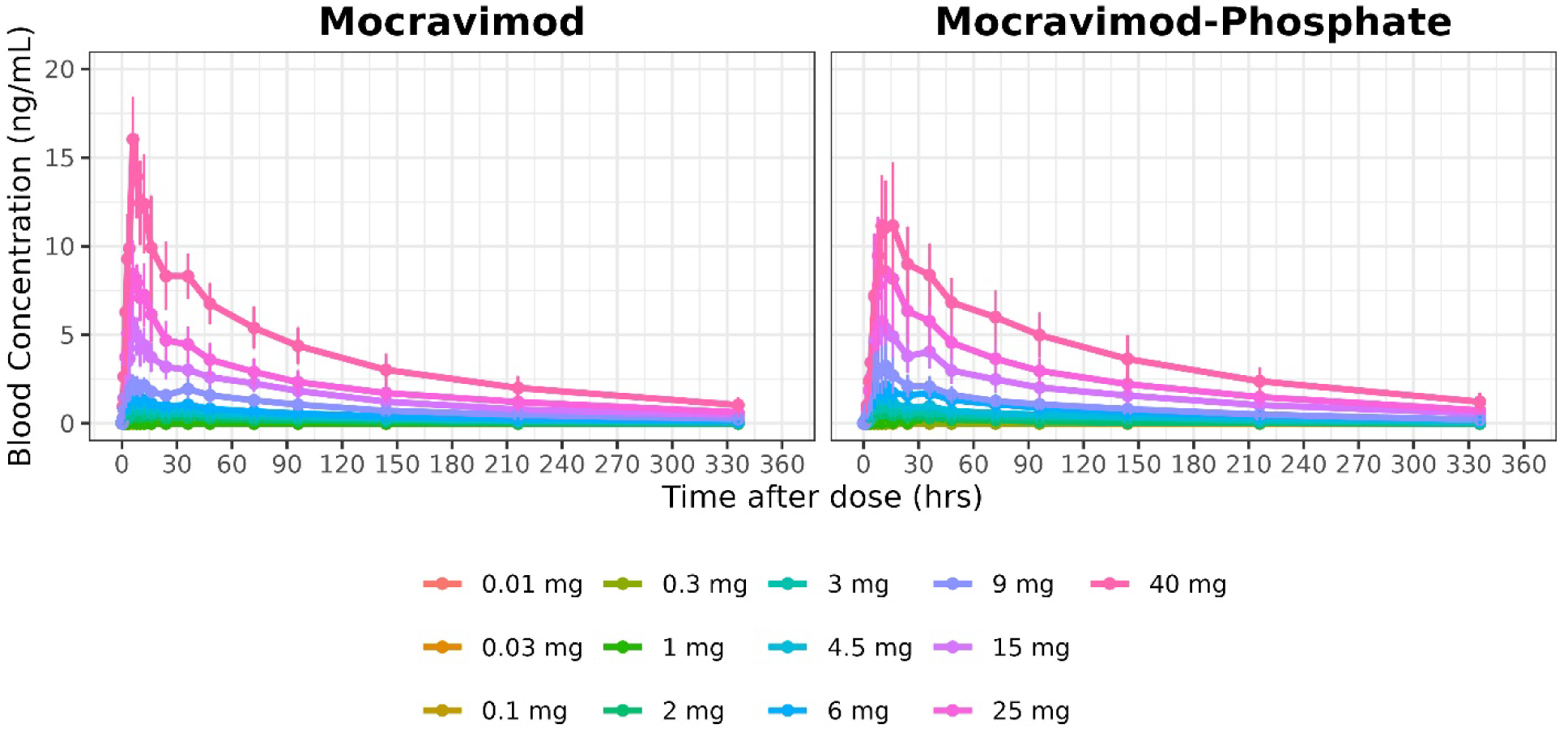
Arithmetic mean concentration-time profiles of mocravimod and mocravimod-phosphate after single dose administration (study part 1)

**Table 2A:** PK parameters of mocravimod (N represents number of subjects dosed in respective group)

| **Mocravimod Dose** | **N** | **AUClast#**<br>**(hr\*ng/mL)** | **AUCinf#**<br>**(hr\*ng/mL)** | **Cmax#**<br>**(ng/mL)** | **T1/2\***<br>**(hr)** | **Tmax\*\***<br>**(hr)** | **Tlag\*\***<br>**(hr)** | **CL/F#**<br>**(L/hr)** | **Vz/F#**<br>**(L)** |
| --- | --- | --- | --- | --- | --- | --- | --- | --- | --- |
| 2 mg | 8 | 32.9 (32%) | - | 0.470<br>(21%) | 90.9 (30.1) | 11.0 (6.02 - 48.1) | 0.375 (0.00 - 1.02) | 25.27 (-) | 3233 (-) |
| 3 mg | 8 | 66.9 (65%) | 182 (26%) | 0.696<br>(28%) | 122 (25.9) | 6.06 (6.03 - 16.1) | 0.392<br>(0.250 - 1.05) | 16.47<br>(26%) | 3148 (16%) |
| 4.5 mg | 13 | 115 (21%) | 118 (22%) | 0.983<br>(15%) | 124 (37.7) | 6.03 (6.00 - 16.1) | 0.250<br>(0.250 - 0.517) | 38.05<br>(22%) | 5814 (27%) |
| 6 mg | 8 | 142 (36%) | 159 (39%) | 1.37 (16%) | 101 (24.6) | 6.08 (6.00 - 12.1) | 0.250 (0.00 - 1.02) | 37.63<br>(39%) | 5332 (17%) |
| 9 mg | 8 | 270 (26%) | 313 (30%) | 2.33 (23%) | 121 (27.8) | 5.97 (5.88 - 36.1) | 0.267<br>(0.200 - 0.550) | 28.78<br>(30%) | 4664 (29%) |
| 15 mg | 8 | 475 (30%) | 559 (37%) | 5.25 (27%) | 132 (33.7) | 6.02 (6.00 - 7.95) | 0.300<br>(0.250 - 0.900) | 26.83<br>(37%) | 4933 (17%) |
| 25 mg | 8 | 686 (24%) | 795 (26%) | 8.73 (17%) | 126 (19.8) | 8.06 (5.93 - 16.2) | 0.258 (0.00 - 0.550) | 31.43<br>(26%) | 5665 (23%) |
| 40 mg | 8 | 1220 (24%) | 1390 (30%) | 15.9 (15%) | 121 (31.9) | 5.99 (5.92 - 6.05) | 0.250 (0.00 - 0.500) | 28.77<br>(30%) | 4629 (21%) |

#Geometric mean (CV% geometric mean)
\*Arithmetic mean (SD) values are presented for T<sub>1/2</sub>
\*\*Median (Min-Max) are presented for T<sub>lag</sub> and T<sub>max</sub>

The active metabolite, mocravimod-phosphate, showed similar PK properties, with slightly higher C_max_ and AUC values and a comparable half-life (Table 2B). Exposure parameters for mocravimod-phosphate (AUC_last_, AUC_inf_, and C_max_) were also more variable than those of mocravimod. This is consistent with a usually higher variability observed with metabolites. T_1/2_ was long and similar to that of mocravimod, without any trend across doses. Except in the 2 mg group where T_1/2_ close to 60 hrs was calculated, mean values were of about 100–135 hrs in the other groups.

**Table 2B:** PK parameters of mocravimod-phosphate (N represents number of subjects dosed in respective group)

| **Mocravimod Dose** | **N** | **AUC<sub>last</sub>#**<br>**(hr\*ng/mL)** | **AUC<sub>inf</sub>#**<br>**(hr\*ng/mL)** | **C<sub>max</sub>#**<br>**(ng/mL)** | **T<sub>1/2</sub>\***<br>**(hr)** | **T<sub>max</sub>\*\***<br>**(hr)** | **T<sub>lag</sub>\*\***<br>**(hr)** | **CL/F#**<br>**(L/hr)** | **V<sub>z</sub>/F#**<br>**(L)** |
| --- | --- | --- | --- | --- | --- | --- | --- | --- | --- |
| 2 mg | 8 | 22.1 (90%) | 38.0 (7%) | 0.630 (47%) | 58.9 (21.7) | 6.07 (5.98 - 12.0) | 2.49 (1.00 - 4.02) | 52.68 (7%) | 2687 (24%) |
| 3 mg | 8 | 70.5 (68%) | - | 0.908 (36%) | 111 (43.1) | 10.0 (6.05 - 16.1) | 1.63 (1.00 - 2.03) | - | - |
| 4.5 mg | 13 | 124 (48%) | 140 (52%) | 1.47 (17%) | 113 (43.6) | 12.0 (6.05 - 36.0) | 1.03 (1.00 - 2.00) | 32.07 (52%) | 4241 (25%) |
| 6 mg | 8 | 215 (51%) | 235 (32%) | 2.15 (35%) | 134 (41.9) | 12.0 (6.05 - 24.1) | 0.767 (0.00 - 1.02) | 25.57 (32%) | 4168 (40%) |
| 9 mg | 8 | 292 (26%) | 323 (25%) | 3.12 (43%) | 101 (17.8) | 11.0 (5.95 - 12.0) | 0.967 (0.467 - 1.08) | 27.84 (25%) | 3992 (31%) |
| 15 mg | 8 | 565 (31%) | 637 (39%) | 6.20 (24%) | 133 (52.6) | 11.0 (6.02 - 16.1) | 0.908 (0.433 - 1.02) | 23.54 (39%) | 3636 (15%) |
| 25 mg | 8 | 834 (29%) | 959 (32%) | 9.03 (32%) | 120 (24.4) | 9.99 (5.92 - 12.0) | 0.483 (0.00 - 1.02) | 26.08 (32%) | 4426 (26%) |
| 40 mg | 8 | 1290 (27%) | 1500 (33%) | 11.9 (26%) | 115 (19.8) | 14.0 (9.90 - 16.2) | 0.508 (0.267 - 1.08) | 26.70 (33%) | 4208 (19%) |

#Geometric mean (CV% geometric mean); \*Arithmetic mean (SD) values are presented for T<sub>1/2</sub>; \*\*Median (Min-Max) are presented for T<sub>lag</sub> and T<sub>max</sub>
AUC: area under the curve; AUC<sub>last</sub>: AUC from zero to last measurable concentration; AUC<sub>inf</sub>: AUC from zero to infinity; CL/F: apparent clearance; C<sub>max</sub>: maximum concentration; CV%: coefficient of variation; T<sub>1/2</sub>: terminal elimination half-life; T<sub>lag</sub>: lag time; T<sub>max</sub>: time to maximum concentration; V<sub>z</sub>/F: apparent volume of distribution

The statistical analysis of the slope estimates and their 90% CI demonstrated lack of dose-proportionality over the entire dose range (2 mg to 40 mg) for AUC_inf_, AUC_last_, or C_max_. Slope estimates were 1.028 (AUC_inf_), 1.166 (AUC_last_), and 1.208 (C_max_), suggesting minor over-proportionality for all parameters. Slope estimates for mocravimod-phosphate were 1.055 (AUC_inf_), 1.255 (AUC_last_), and 1.027 (C_max_), suggesting the same as mocravimod, a minor over-proportionality for all parameters.

### Food effect results

PK parameters confirmed the observations made from the concentration-time curves (Figure 3), suggesting that food had a very limited effect on both mocravimod and mocravimod-phosphate distribution, but with opposite trends, i.e. higher C_max_ (+12%) and lower T_max_ (6 versus 11 hrs) for mocravimod, and lower C_max_ (–20%) and higher T_max_ (12 versus 10 hrs) for mocravimod-phosphate. T_1/2_ seemed to increase with food for mocravimod (+8.5%), whereas it seemed to decrease for mocravimod-phosphate (–14%) (Table 3 and Table S1). The 90% CIs for AUC_last_ and AUC_inf_ of mocravimod were fully within the 0.80–1.25 range, indicating a lack of food effect. The 90% CI for C_max_ slightly exceeded the upper bound and did not include 1, so a small food-related increase in C_max_ could not be ruled out. For mocravimod-phosphate, the AUC_last_ CI met the bioequivalence range, but the AUC_inf_ CI did not, although it included 1 and the ratio (1.05) was close to unity. The C_max_ CI was only partly within range and overlapped the lower bound, suggesting a possible food-related decrease in C_max_.

**Figure 3:**
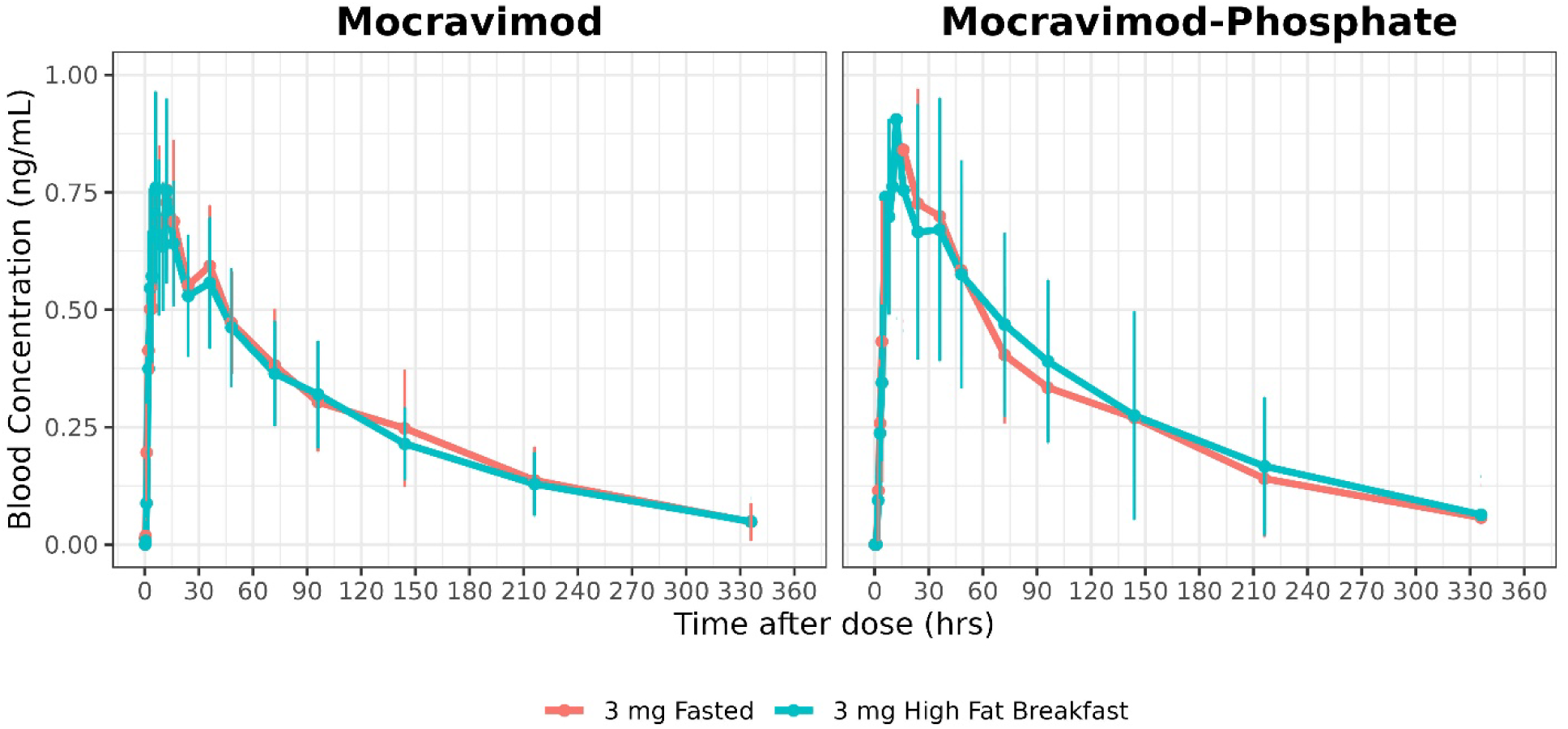
Arithmetic mean concentration-time profiles of mocravimod and mocravimod-phosphate after 3 mg high fed breakfast or 3 mg fasted (study part 2)

**Table 3:** Geometric Mean Ratio Results of the Food Effect (Part 2)

| Analyte | Parameter | Adjusted GM* |  | GMR* |  |  |
| --- | --- | --- | --- | --- | --- | --- |
|  |  | Fed | Fasted | Estimate<br>Fed/Fasted | Lower<br>90%<br>CL | Upper<br>90%<br>CL |
| Mocravimod | AUClast<br>(hr*ng/mL) | 74.8 | 77.0 | 0.97 | 0.89 | 1.06 |
| Mocravimod | AUCinf<br>(hr*ng/mL) | 88.1 | 86.0 | 1.02 | 0.93 | 1.13 |
| Mocravimod | Cmax<br>(ng/mL) | 0.877 | 0.744 | 1.18 | 1.06 | 1.31 |
| Mocravimod-phosphate | AUClast<br>(hr*ng/mL) | 81.7 | 85.0 | 0.96 | 0.86 | 1.07 |
| Mocravimod-phosphate | AUCinf<br>(hr*ng/mL) | 124 | 119 | 1.05 | 0.68 | 1.62 |
| Mocravimod-phosphate | Cmax<br>(ng/mL) | 0.922 | 1.11 | 0.83 | 0.69 | 1.00 |

\*
AUC: area under the curve; AUClast: AUC from zero to last measurable concentration; AUC<sub>inf</sub>: AUC from zero to infinity; CL: confidence limit; C<sub>max</sub>: maximum concentration; GM: geometric mean; GMR: geometric mean ratio

### Pharmacodynamic Results

#### Peripheral lymphopenia

Mocravimod and mocravimod-phosphate induced a dose-dependent reduction in absolute lymphocyte counts (Figure 4). Significant decreases were observed at doses ≥2 mg, with the maximum mean percentage decrease occurring between 8 and 24 hrs post-dose (Figure S2). The mean maximum decrease ranged from approximately 8% (0.01 mg) to 87% (25 mg). For the pooled placebo group, the mean maximum decrease was 4.3%. At doses ≥2 mg, all participants experienced significant lymphocyte reduction, with recovery to ≥1000/μL by end of study in all participants up to 9 mg; recovery was slower at higher doses, with only 12.5% of participants recovering following a dose of 40 mg.

**Figure 4:**
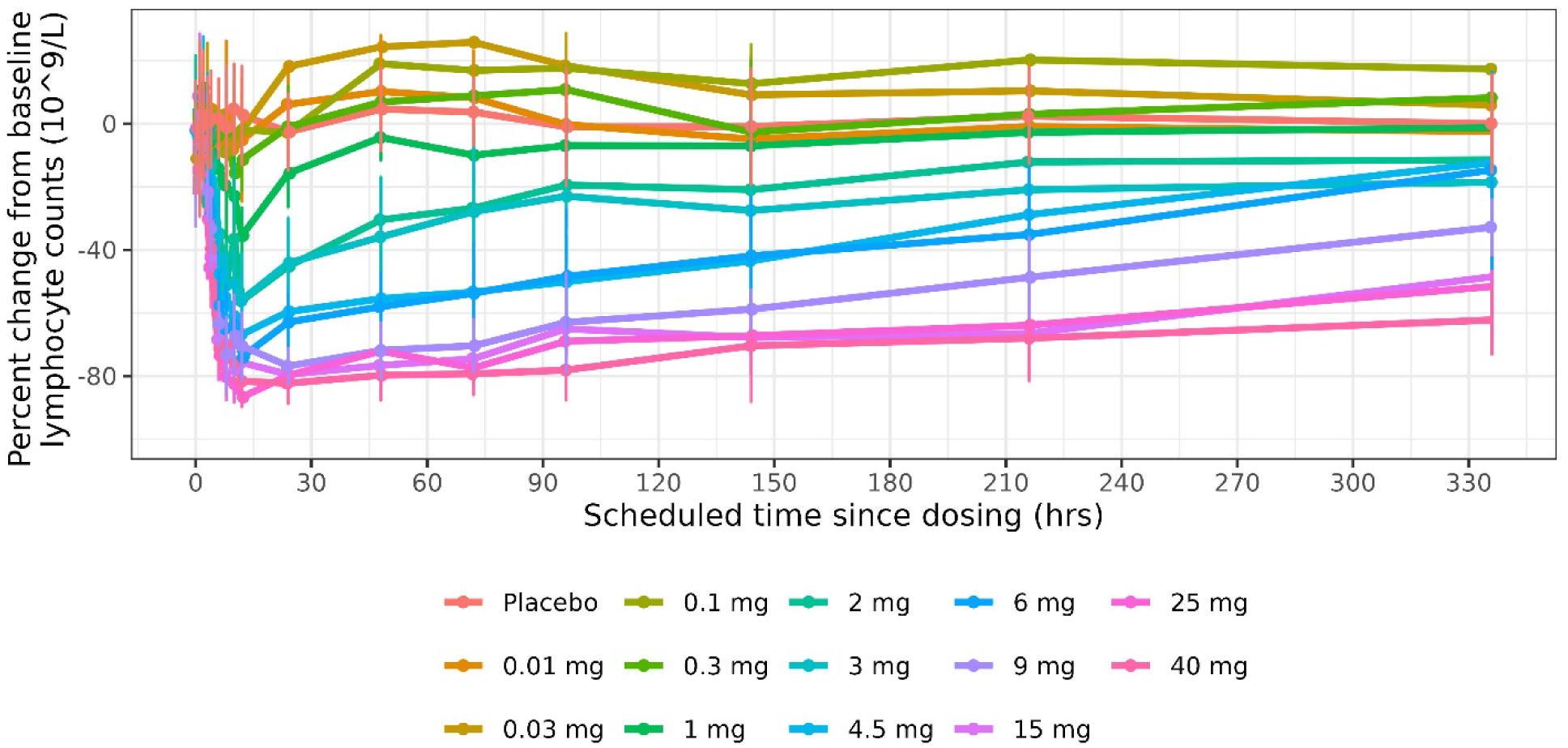
Percent change in lymphocyte counts over time by dose group.

The mocravimod doses were compared to placebo with respect to the log transformed ratios for E_max_ and AUC_0-24_ of the absolute lymphocyte counts (ALC) (Table S2). For ALC E_max_ differences relative to placebo were statistically significant at mocravimod doses of 2 mg and above, whereas for ALC AUC_0-24_ statistical significance was reached at mocravimod doses of 1 mg and above.

### General Safety Results

A total of 76 of the 136 participants who received mocravimod (55.9%) reported adverse events (AEs). Of the 27 participants who received placebo, 10 (37%) experienced AEs. There were no serious adverse events (SAEs) or deaths during this study. A summary of adverse events is presented in Table 4.

**Table 4:**
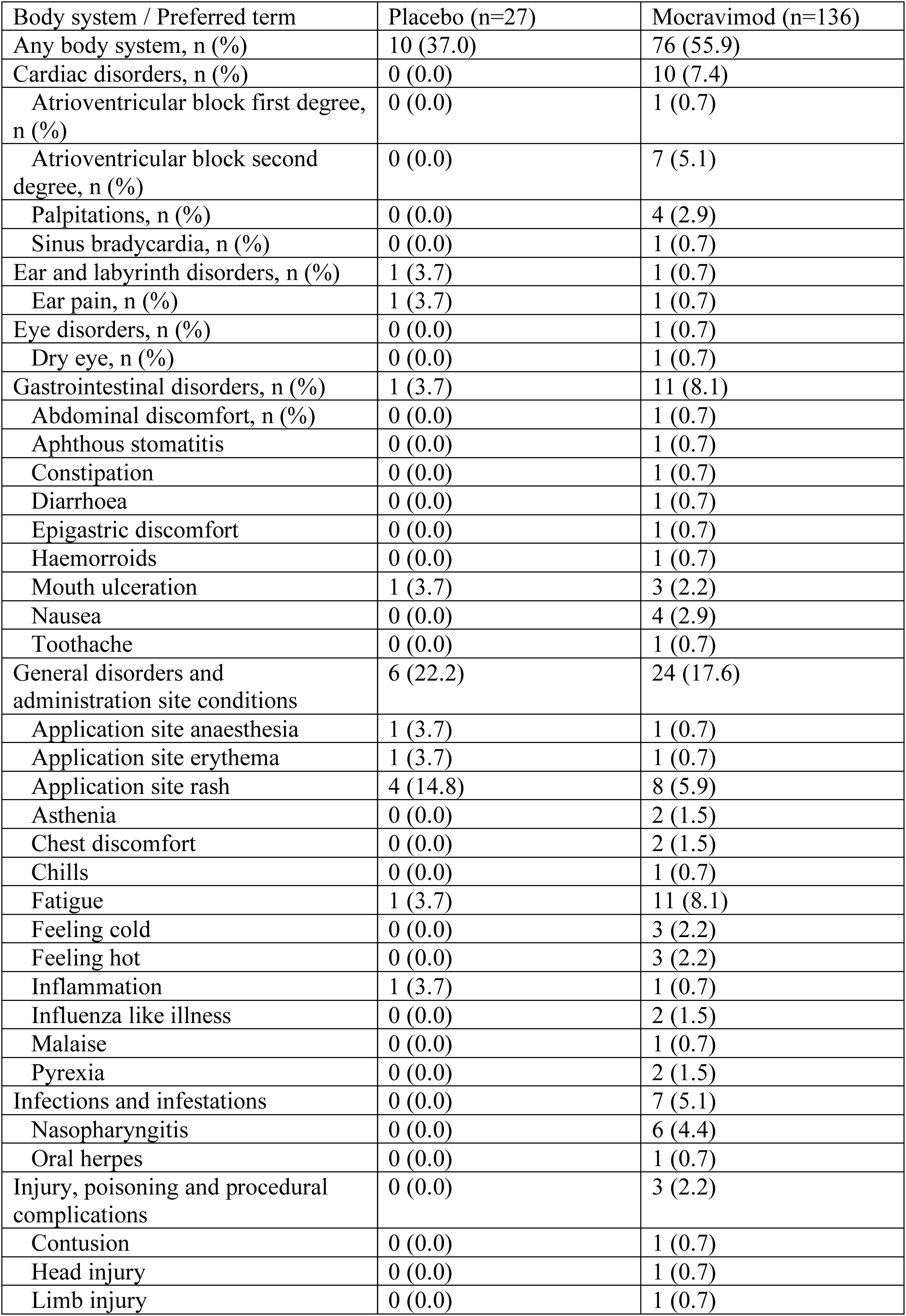
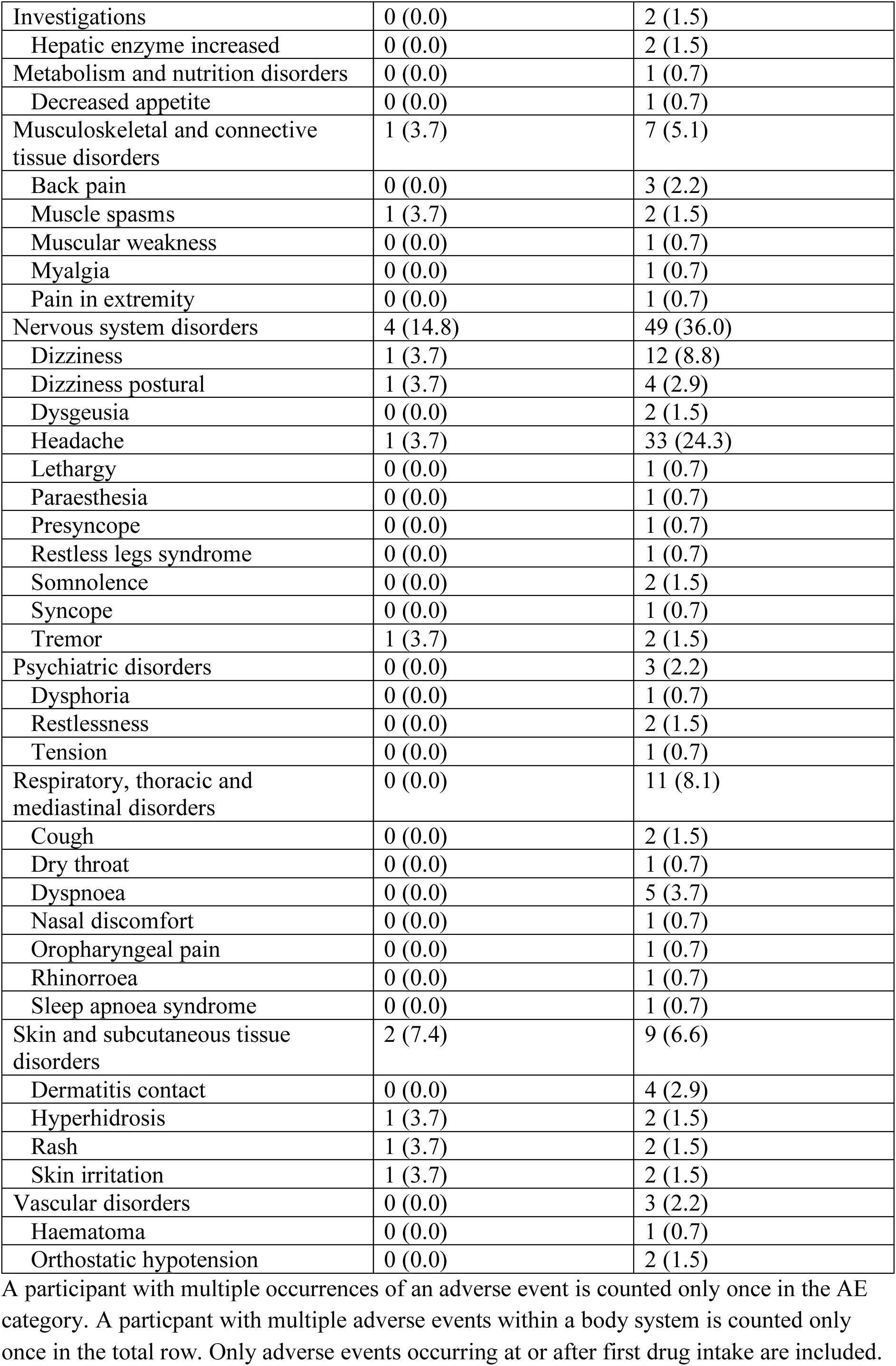
Safety evaluation - Participants with adverse events by body system and preferred term – Safety population.

The AEs were mostly transient and either mild or moderate in severity. In participants receiving mocravimod, the most frequently affected system organ classes (SOCs), which involved ≥5% of participants, were nervous system disorders (n=49, 36%), general disorders and administration site conditions (n=24, 17.6%), gastrointestinal disorders (n=11, 8.1%), respiratory, thoracic and mediastinal disorders (n=11, 8.1%), skin and subcutaneous tissue disorders (n=9, 6.6%), infections and infestations (n=7, 5.1%), and musculoskeletal and connective tissue disorders (n=7, 5.1%). The most common adverse events (AEs) were headache (n=33, 24.3%), dizziness (n=12, 8.8%), and fatigue (n=11, 8.1%) and atrioventricular (AV) block second degree (n=7, 5.1%), all of which were mild or moderate and transient.

Gastrointestinal, respiratory, and skin-related AEs were infrequent and not dose-dependent. Laboratory assessments revealed sporadic, non-clinically significant abnormalities, except for two cases of transient, mild liver enzyme elevations (one suspected related to study drug) (Table S4). No clinically significant changes in pulmonary function were observed.

### Cardiac Safety

Cardiac AEs were reported by 10 participants: second degree atrioventricular block (7 subjects, 5.1%) was reported in 6 mg, 9 mg, 15 mg, 25 mg and 40 mg mocravimod dose groups, palpitations (4 subjects, 2.9%) were reported in the 2 mg, 4.5 mg and 6 mg mocravimod dose groups, first degree atrioventricular block (1 subject, 0.7%) was reported in 9 mg mocravimod dose group; and sinus bradycardia (1 subject, 0.7%) was reported in the 40 mg mocravimod dose group.

The mean hourly heart rate data for Day −1 and Day 1 are shown in Figure 5. At baseline (Day −1), mean hourly heart rate profiles were comparable across all treatment groups, with no notable differences observed throughout the day. On Day 1, the mean hourly heart rate profiles for mocravimod dose groups of ≥1 mg differed from those of the placebo group. A rapid reduction in mean hourly heart rate relative to placebo was observed within the first 6 hrs post-dose and persisted for the remainder of the day (Figure S3). The reduction in heart rate was not dose dependent as most pronounced between 0.3 mg and 2 mg, and there was no suggestion that heart rate was reduced further above a dose of 2 mg (Table S3).

**Figure 5:**
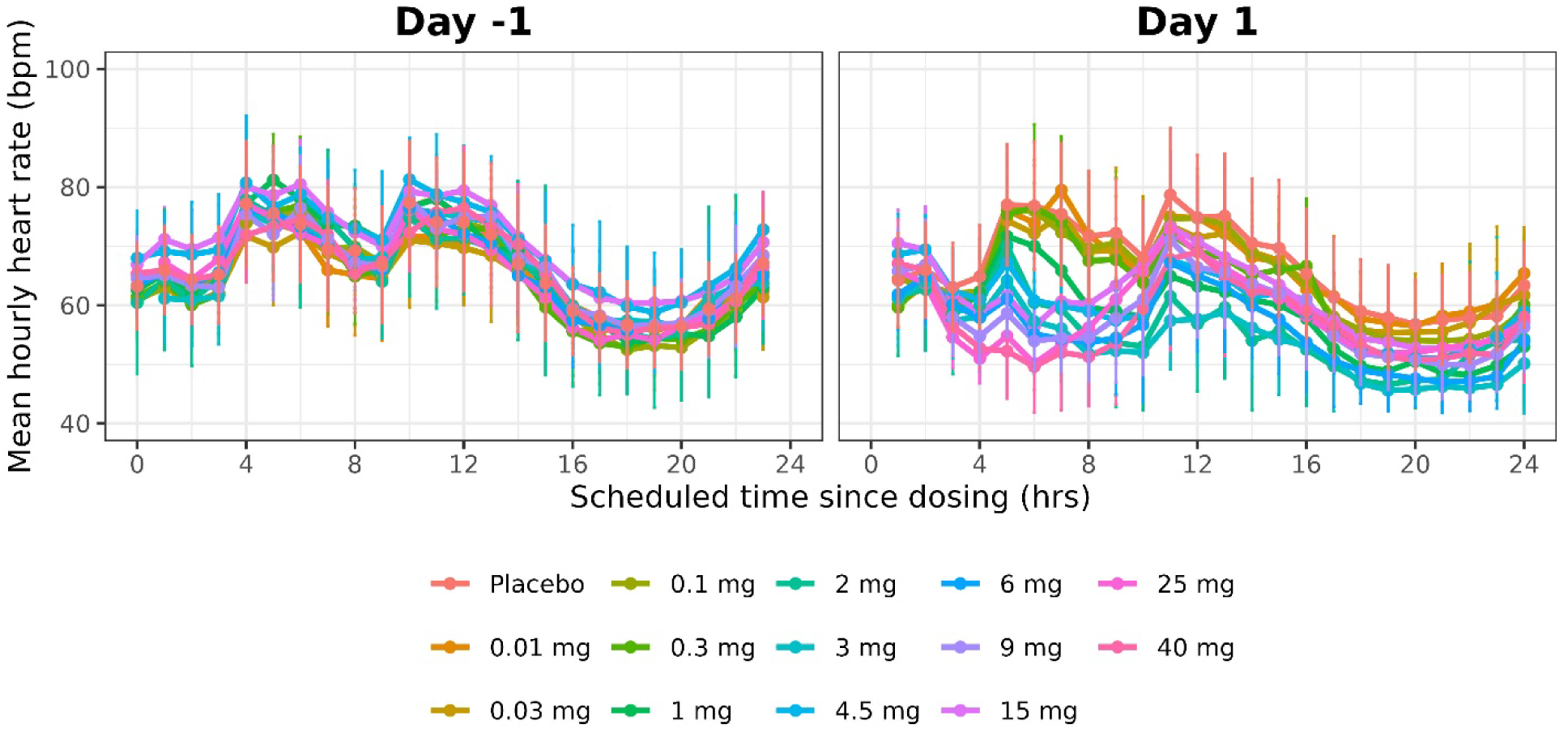
Mean hourly heart rate per dose group. Day -1 (baseline) and Day 1 (treatment).

Atrioventricular (AV) block (Mobitz I), mostly second degree, was observed at every dose level from 6 mg onwards and seemed independent of dose received. These events were transient, asymptomatic, and not deemed clinically significant (Table S4). Four cases of second-degree AV block (Mobitz I/Wenckebach) were considered possibly related to mocravimod: one at 6 mg, one at 15 mg, and two at 40 mg (all occurring on Day 1). Despite these conduction events, QRS changes were not significant, QTcF showed variable changes with no clear dose- or day-relationship, and there were no serious arrhythmias, deaths, or serious adverse events during the study.

## Discussion

In this first-in-human study, mocravimod, an S1P receptor modulator that binds to four of the five S1P receptor isoforms, with the highest potency for isoforms 1 and 5, demonstrated predictable, dose-proportional pharmacokinetics, robust pharmacodynamic effects on lymphocyte counts, and a favourable safety profile in healthy volunteers.

The PK profile is characterised by slow absorption, dose-proportional exposure, extensive distribution, and a long terminal half-life. Mocravimod, as a precursor to the active moiety mocravimod-phosphate, allows to obtain mocravimod-phosphate at approximately equimolar circulating levels. Apparent clearance (CL/F) was variable, fluctuating between 16 and 38 L/hr for mocravimod and 16 and 53 L/hr for mocravimod phosphate (Table 2), and within the range of the hepatic blood flow (87 L/hr) in healthy participants [16]. The volume of distribution of 3000–6000 L was much larger than the total body water (42 L) of healthy participants, which suggests extensive tissue distribution. The administration of 3 mg mocravimod under fed and fasted conditions revealed minimal impact of food on overall exposure (AUC_inf_ and AUC_last_) and a modest effect on C_max_ so that mocravimod can be administered without regard to food.

Although dose-proportionality was a deviation from statistical significance, such deviations generally have only a minor impact on mocravimod exposure when considering small increases in dose factor (from 2-fold to 5-fold). Mocravimod’s PK profile—slow absorption, long half-life, and dose-proportional exposure—mirrors other S1P receptor modulators such as fingolimod and siponimod, though the half-life of mocravimod (90–130 hrs) is at the upper end of the range reported for this class [17]. While the observed PK profile suggests the potential for once-daily or less frequent dosing, accumulation and steady-state effects require further investigation. Recent drug-drug interaction studies demonstrated that a single dose of 3 mg of mocravimod did not show a clinically relevant interaction with multiple dosing of itraconazole or cyclosporin in healthy volunteers [18].

Pharmacodynamically, mocravimod induces robust, dose-dependent reductions in peripheral lymphocyte counts, consistent with its mechanism of S1P1 antagonism. The results align with the established pharmacology of S1P modulators. Fingolimod, the first-in-class agent, and newer agents such as siponimod, ozanimod, and ponesimod, all induce reversible lymphopenia by sequestering lymphocytes in secondary lymphoid organs [1, 2]. The observed peripheral lymphopenia is also comparable, with maximal decreases of 70–90% at higher doses, and recovery to baseline within weeks after cessation, as seen with other S1P receptor modulators.

Transient bradycardia is a well-known class effect of S1P1 receptor modulators. However, this bradycardia typically normalizes with repeated dosing due to internalization of S1P1/S1P3 receptors on cardiac cells. S1P1 modulators with short half-lives such as ponesimod and ozanimod require up-titration to reduce bradycardia, and the initial HR decreases with up-titrated ponesimod 2.5 mg (8 bpm) and ozanimod 0.3 mg (7 bpm) are comparable to mocravimod [19].

Mocravimod was generally well tolerated up to the maximum tested dose of 40 mg. At 40 mg the stopping criteria for further dose escalation was met as one participant on active treatment developed symptomatic bradycardia. The safety profile is favourable, with adverse events predominantly mild or moderate, and class-typical cardiovascular effects (transient bradycardia, mostly asymptomatic AV blocks) observed only at higher doses at least 6 mg and without clinical sequelae. The safety profile was consistent with the known class effects of S1P receptor modulators, including reversible lymphopenia and transient bradycardia. No clinically significant pulmonary, neurological, or laboratory safety signals were identified. The overall incidence and severity of AEs did not increase substantially with dose, except for a higher frequency of benign cardiac conduction abnormalities at the highest doses. Cardiovascular effects, particularly transient bradycardia and AV conduction delays, are well-documented class effects, attributed to S1P1 receptor modulation in cardiac tissue [4]. In this study, bradycardia and second-degree AV block were observed at doses ≥6 mg but were asymptomatic and resolved spontaneously. No clinically significant QT prolongation or arrhythmias were detected, and no dose-limiting toxicities occurred. These findings are consistent with the safety profiles of approved S1P receptor modulators, where first-dose cardiac monitoring is standard for higher-risk patients [3].

Mocravimod’s strong affinity for S1P1 and S1P5, and its reduced CNS penetration compared to fingolimod, may offer advantages in terms of neuropsychiatric safety and washout kinetics. Recent reviews highlight the need for S1P receptor modulators with improved selectivity and safety, particularly for use in populations at risk for CNS or cardiac complications [20].

The immunomodulatory effects of S1P receptor modulators have been leveraged primarily in autoimmune diseases, but there is a substantial medical need in their application to haematological malignancies and transplantation. In the context of allo-HCT for AML, the major challenge is to prevent GvHD while preserving or enhancing the GvL effect [8, 21]. S1P receptor modulators, by restricting T-cell egress from lymphoid tissues, can reduce the migration of alloreactive T cells to GvHD target organs, potentially mitigating the risk of GvHD without impairing anti-leukaemic immunity [7, 13]. Preclinical studies of mocravimod and other S1P receptor modulators have shown that this approach can “decouple” GvHD from GvL, improving survival in murine models of allo-HCT. Early-phase clinical trials of mocravimod in patients undergoing allo-HCT for AML have reported encouraging safety and efficacy signals, with trends toward reduced relapse and improved survival compared to historical controls. The ongoing MO-TRANS Phase 3 trial (NCT05429632) will provide evidence for this strategy [22].

## Conclusion

The observed safety and tolerability in this first in human single ascending dose study, including manageable cardiovascular effects and absence of serious toxicity, support further clinical development of mocravimod in immune-mediated and haematological diseases.

## Data Availability

Data produced in the present study are available upon reasonable request to the authors.

## Acknowledgements

The authors wish to thank Dr. Margit Hemetsberger, Vienna, Austria, for editorial services, funded by Priothera SAS, Saint-Louis, France. This study was conducted by Novartis Pharma AG, Basel, Switzerland.

## Author contributions

All authors have substantially contributed to the conception of the work; or the acquisition, analysis, or interpretation of data for the work. DH has drafted the work; DK, JH reviewed it critically for important intellectual content. All authors have approved the final version of the work to be published and agree to be accountable for all aspects of the work in ensuring that questions related to the accuracy or integrity of any part of the work are appropriately investigated and resolved.

