## Supplemental figures and tables for "A Randomized, Double-Blind, Placebo-Controlled, Single Ascending Oral Dose Study of Mocravimod: Safety, Tolerability, Pharmacokinetics, and Pharmacodynamics in Healthy Participants"

**Supplemental materials**

**Figure S1: Arithmetic mean concentration-time profiles of mocravimod and mocravimod-phosphate after single dose administration (study part 1, 2-40 mg) on logarithmic scale (y-axis)**

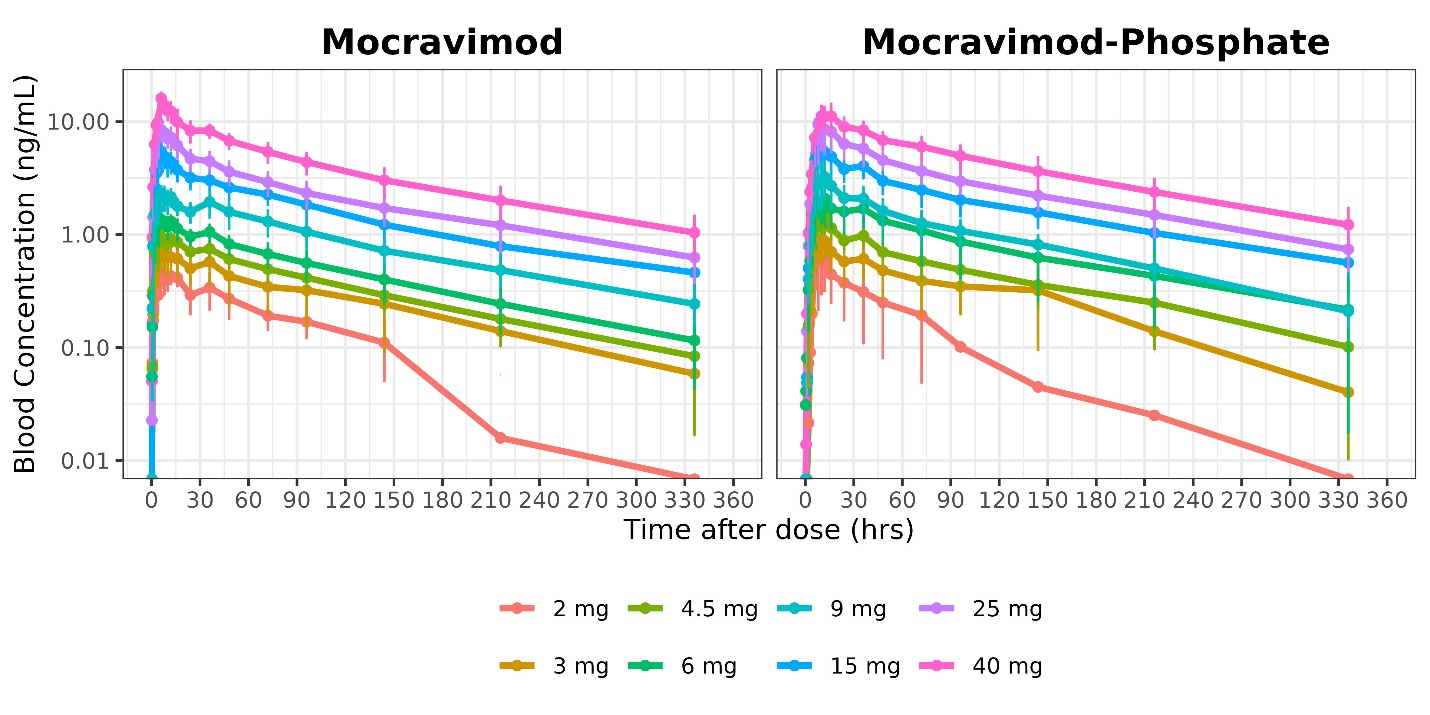

**Figure S2: Percent change in lymphocyte counts per dose group on Day 1 after dosing (study part 1)**

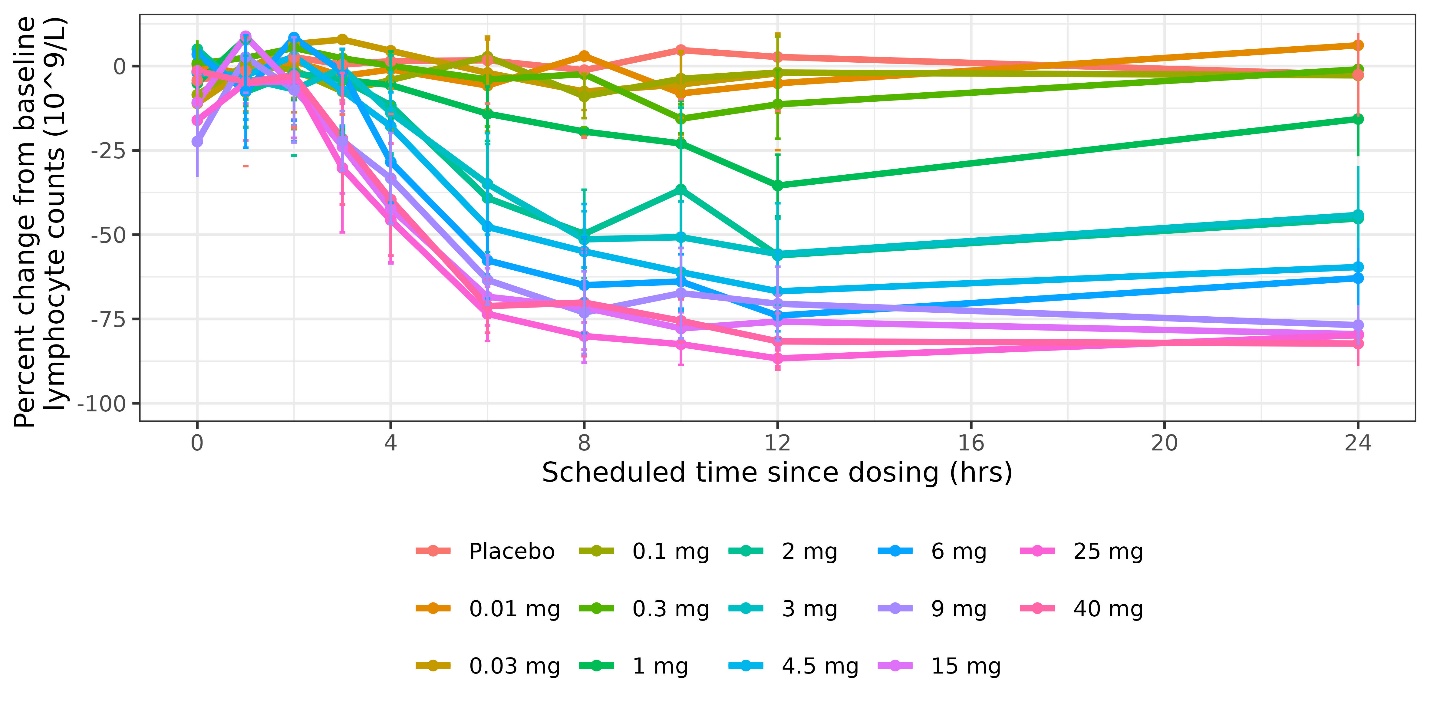

**Figure S3: Mean change in hourly heart rate per dose group (study part 1)**

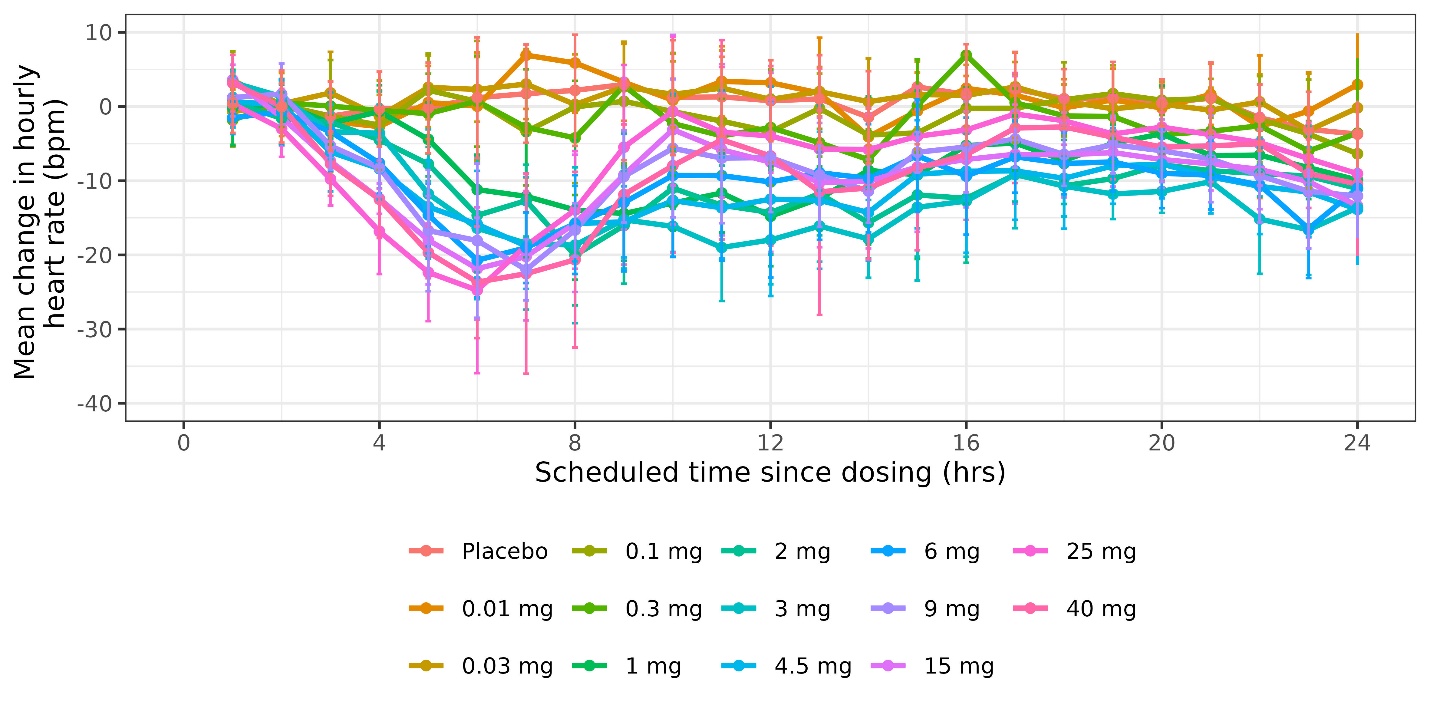

**Table S1: Mocravimod and mocravimod-phosphate PK parameters in fasted and fed condition after a single dose of 3 mg (study part 2)**

|  | **Mocravimod** | | **Mocravimod-Phosphate** | |
| --- | --- | --- | --- | --- |
|  | **Fasted N=8** | **Fed N=9** | **Fasted N=8** | **Fed N=9** |
| AUCinf (hr*ng/mL) | 88.0 (43%) | 86.5 (43%) | 126 (56%) | 122 (53%) |
| AUClast (hr*ng/mL) | 79.0 (44%) | 73.4 (42%) | 84.1 (59%) | 79.6 (75%) |
| Cmax (ng/mL) | 0.777 (23%) | 0.871 (23%) | 1.14 (50%) | 0.908 (40%) |
| T1/2 (hr) | 88.6 (23.2) | 96.1 (22.1) | 109 (46.7) | 93.8 (31.8) |
| Tmax (hr) | 11.0 (6.00–12.0) | 6.02 (2.98–12.1) | 9.99 (6.00–12.0) | 12.0 (8.02–24.0) |

AUC: area under the curve; AUClast: AUC from zero to last measurable concentration; AUCinf: AUC from zero to infinity; Cmax: maximum concentration; CV%: coefficient of variation; T1/2; terminal elimination half-life; Tmax: time to maximum concentration

Geometric mean (CV% geometric mean) are presented for AUCinf, AUClast, and Cmax. Arithmetic mean (SD) are presented for T1/2. Median (Min-Max) are presented for Tmax.

**Table S2: Geometric mean ratio for treatment versus placebo for lymphocyte parameters**

| **Dose group** | **GMR Emax (10^9/L)** | **95% CI** | **p-value** | **GMR AUC0-24 (hr*10^9/L)** | **95% CI** | **p-value** |
| --- | --- | --- | --- | --- | --- | --- |
| 0.01 mg | 0.98 | (0.80 - 1.19) | 0.8035 | 0.95 | (0.86 - 1.06) | 0.3812 |
| 0.03 mg | 1.04 | (0.86 - 1.27) | 0.6775 | 1.03 | (0.93 - 1.14) | 0.5813 |
| 0.1 mg | 1.02 | (0.84 - 1.24) | 0.8403 | 0.97 | (0.88 - 1.07) | 0.5557 |
| 0.3 mg | 1.01 | (0.83 - 1.23) | 0.9185 | 0.95 | (0.85 - 1.05) | 0.2709 |
| 1 mg | 0.87 | (0.71 - 1.06) | 0.1560 | 0.80 | (0.72 - 0.89) | <0.0001 |
| 2 mg | 0.55 | 0.45 - 0.67) | <0.0001 | 0.59 | (0.54 - 0.66) | <0.0001 |
| 3 mg | 0.52 | (0.43 - 0.63) | <0.0001 | 0.59 | (0.53 - 0.65) | <0.0001 |
| 4.5 mg | 0.39 | (0.33 - 0.46) | <0.0001 | 0.49 | (0.45 - 0.53) | <0.0001 |
| 6 mg | 0.33 | (0.27 - 0.40) | <0.0001 | 0.45 | (0.40 - 0.50) | <0.0001 |
| 9 mg | 0.26 | (0.21 - 0.31 | <0.0001 | 0.39 | (0.35 - 0.43) | <0.0001 |
| 15 mg | 0.24 | (0.19 - 0.29) | <0.0001 | 0.35 | (0.31 - 0.39) | <0.0001 |
| 25 mg | 0.16 | (0.14 - 0.20) | <0.0001 | 0.29 | (0.26 - 0.32) | <0.0001 |
| 40 mg | 0.18 | (0.15 - 0.22) | <0.0001 | 0.33 | (0.30 - 0.36) | <0.0001 |

AUC: area under the curve; CI: confidence interval; Emax: maximum effect; GMR: geometric mean ratio

**Table S3: Geometric mean ratio for treatment versus placebo heart rate parameters**

| **Dose group** | **GMR Emax (mmHg)** | **95% CI** | **p-value** | **GMR AUC0-24 (min*bpm)** | **95% CI** | **p-value** |
| --- | --- | --- | --- | --- | --- | --- |
| 0.01 mg | 1.01 | (0.93–1.09) | 0.8406 | 1.01 | (0.96 - 1.06) | 0.7665 |
| 0.03 mg | 1.01 | (0.94–1.09) | 0.7028 | 1.00 | (0.96 - 1.05) | 0.8657 |
| 0.1 mg | 1.01 | (0.94–1.09) | 0.7222 | 0.98 | (0.94 - 1.02) | 0.3424 |
| 0.3 mg | 0.96 | (0.89–1.03) | 0.2306 | 0.97 | (0.92 - 1.01) | 0.1118 |
| 1 mg | 0.91 | (0.85–0.98) | 0.0119 | 0.88 | (0.84 - 0.92) | <.0001 |
| 2 mg | 0.83 | (0.77–0.90) | <.0001 | 0.82 | (0.79 - 0.87) | <.0001 |
| 3 mg | 0.70 | (0.64–0.77) | <.0001 | 0.83 | (0.79 - 0.88) | <.0001 |
| 4.5 mg | 0.87 | (0.81–0.93) | 0.0002 | 0.84 | (0.81 - 0.88) | <.0001 |
| 6 mg | 0.84 | (0.79–0.91) | <.0001 | 0.84 | (0.80 - 0.88) | <.0001 |
| 9 mg | 0.90 | (0.84–0.97) | 0.0041 | 0.86 | (0.83 - 0.90) | <.0001 |
| 15 mg | 0.87 | (0.80–0.94) | 0.0004 | 0.86 | (0.82 - 0.90) | <.0001 |
| 25 mg | 0.85 | (0.77–0.93) | 0.0008 | 0.89 | (0.84 - 0.95) | 0.0002 |
| 40 mg | 0.84 | (0.79–0.91) | <.0001 | 0.85 | (0.82 - 0.89) | <.0001 |

AUC: area under the curve; CI, confidence interval; Emax: maximum effect; GMR: geometric mean ratio

**Table S4. Subjects with adverse events by body system and preferred term – Safety population**

| **Body system/ Preferred Term** | **Placebo** | **0.01 mg** | **0.03 mg** | **0.1 mg** | **0.3 mg** | **1 mg** | **2 mg** | **3 mg** | **4.5 mg** | **6 mg** | **9 mg** | **15 mg** | **25 mg** | **40 mg** | **Total** |
| --- | --- | --- | --- | --- | --- | --- | --- | --- | --- | --- | --- | --- | --- | --- | --- |
|  | N=27 | N=8 | N=8 | N=8 | N=8 | N=8 | N=8 | N=8 | N=13 | N=8 | N=8 | N=8 | N=8 | N=8 | N=136 |
| **Any body system -Total** | **10 (37.0)** | **3 (37.5)** | **5 (62.5)** | **7 (87.5)** | **5 (62.5)** | **2 (25.0)** | **1 (12.5)** | **1 (12.5)** | **10 (76.9)** | **5 (62.5)** | **6 (75.0)** | **5 (62.5)** | **8 (100)** | **8 (100)** | **76 (55.9)** |
| **Cardiac disorders - Total** | **0 (0.0)** | **0 (0.0)** | **0 (0.0)** | **0 (0.0)** | **0 (0.0)** | **0 (0.0)** | **1 (12.5)** | **0 (0.0)** | **2 (15.4)** | **2 (25.0)** | **1 (12.5)** | **1 (12.5)** | **1 (12.5)** | **2 (25.0)** | **10 (7.4)** |
| Atrioventricular block first degree | 0 (0.0) | 0 (0.0) | 0 (0.0) | 0 (0.0) | 0 (0.0) | 0 (0.0) | 0 (0.0) | 0 (0.0) | 0 (0.0) | 0 (0.0) | 1 (12.5) | 0 (0.0) | 0 (0.0) | 0 (0.0) | 1 (0.7) |
| Atrioventricular block second degree | 0 (0.0) | 0 (0.0) | 0 (0.0) | 0 (0.0) | 0 (0.0) | 0 (0.0) | 0 (0.0) | 0 (0.0) | 0 (0.0) | 2 (25.0) | 1 (12.5) | 1 (12.5) | 1 (12.5) | 2 (25.0) | 7 (5.1) |
| Palpitations | 0 (0.0) | 0 (0.0) | 0 (0.0) | 0 (0.0) | 0 (0.0) | 0 (0.0) | 1 (12.5) | 0 (0.0) | 2 (15.4) | 1 (12.5) | 0 (0.0) | 0 (0.0) | 0 (0.0) | 0 (0.0) | 4 (2.9) |
| Sinus bradycardia | 0 (0.0) | 0 (0.0) | 0 (0.0) | 0 (0.0) | 0 (0.0) | 0 (0.0) | 0 (0.0) | 0 (0.0) | 0 (0.0) | 0 (0.0) | 0 (0.0) | 0 (0.0) | 0 (0.0) | 1 (12.5) | 1 (0.7) |
| **Ear and labyrinth disorders - Total** | **1 (3.7)** | **0 (0.0)** | **0 (0.0)** | **0 (0.0)** | **0 (0.0)** | **0 (0.0)** | **0 (0.0)** | **0 (0.0)** | **0 (0.0)** | **0 (0.0)** | **0 (0.0)** | **0 (0.0)** | **0 (0.0)** | **0 (0.0)** | **1 (0.7)** |
| Ear pain | 1 (3.7) | 0 (0.0) | 0 (0.0) | 0 (0.0) | 0 (0.0) | 0 (0.0) | 0 (0.0) | 0 (0.0) | 0 (0.0) | 0 (0.0) | 0 (0.0) | 0 (0.0) | 0 (0.0) | 0 (0.0) | 1 (0.7) |
| Eye disorders - Total | 0 (0.0) | 0 (0.0) | 0 (0.0) | 0 (0.0) | 0 (0.0) | 0 (0.0) | 0 (0.0) | 0 (0.0) | 0 (0.0) | 0 (0.0) | 0 (0.0) | 0 (0.0) | 1 (12.5) | 0 (0.0) | 1 (0.7) |
| Dry eye | 0 (0.0) | 0 (0.0) | 0 (0.0) | 0 (0.0) | 0 (0.0) | 0 (0.0) | 0 (0.0) | 0 (0.0) | 0 (0.0) | 0 (0.0) | 0 (0.0) | 0 (0.0) | 1 (12.5) | 0 (0.0) | 1 (0.7) |
| **Gastrointestinal disorders - Total** | **1 (3.7)** | **1 (12.5)** | **0 (0.0)** | **0 (0.0)** | **0 (0.0)** | **0 (0.0)** | **0 (0.0)** | **0 (0.0)** | **2 (15.4)** | **2 (25.0)** | **0 (0.0)** | **1 (12.5)** | **2 (25.0)** | **2 (25.0)** | **11 (8.1)** |
| Abdominal discomfort | 0 (0.0) | 0 (0.0) | 0 (0.0) | 0 (0.0) | 0 (0.0) | 0 (0.0) | 0 (0.0) | 0 (0.0) | 0 (0.0) | 1 (12.5) | 0 (0.0) | 0 (0.0) | 0 (0.0) | 0 (0.0) | 1 (0.7) |
| Aphthous stomatitis | 0 (0.0) | 0 (0.0) | 0 (0.0) | 0 (0.0) | 0 (0.0) | 0 (0.0) | 0 (0.0) | 0 (0.0) | 0 (0.0) | 1 (12.5) | 0 (0.0) | 0 (0.0) | 0 (0.0) | 0 (0.0) | 1 (0.7) |
| Constipation | 0 (0.0) | 0 (0.0) | 0 (0.0) | 0 (0.0) | 0 (0.0) | 0 (0.0) | 0 (0.0) | 0 (0.0) | 1 (7.7) | 0 (0.0) | 0 (0.0) | 0 (0.0) | 0 (0.0) | 0 (0.0) | 1 (0.7) |
| Diarrhoea | 0 (0.0) | 0 (0.0) | 0 (0.0) | 0 (0.0) | 0 (0.0) | 0 (0.0) | 0 (0.0) | 0 (0.0) | 0 (0.0) | 1 (12.5) | 0 (0.0) | 0 (0.0) | 0 (0.0) | 0 (0.0) | 1 (0.7) |
| Epigastric discomfort | 0 (0.0) | 0 (0.0) | 0 (0.0) | 0 (0.0) | 0 (0.0) | 0 (0.0) | 0 (0.0) | 0 (0.0) | 0 (0.0) | 0 (0.0) | 0 (0.0) | 0 (0.0) | 1 (12.5) | 0 (0.0) | 1 (0.7) |
| Haemorrhoids | 0 (0.0) | 0 (0.0) | 0 (0.0) | 0 (0.0) | 0 (0.0) | 0 (0.0) | 0 (0.0) | 0 (0.0) | 1 (7.7) | 0 (0.0) | 0 (0.0) | 0 (0.0) | 0 (0.0) | 0 (0.0) | 1 (0.7) |
| Mouth ulceration | 1 (3.7) | 0 (0.0) | 0 (0.0) | 0 (0.0) | 0 (0.0) | 0 (0.0) | 0 (0.0) | 0 (0.0) | 0 (0.0) | 0 (0.0) | 0 (0.0) | 1 (12.5) | 0 (0.0) | 1 (12.5) | 3 (2.2) |
| Nausea | 0 (0.0) | 0 (0.0) | 0 (0.0) | 0 (0.0) | 0 (0.0) | 0 (0.0) | 0 (0.0) | 0 (0.0) | 0 (0.0) | 1 (12.5) | 0 (0.0) | 0 (0.0) | 1 (12.5) | 2 (25.0) | 4 (2.9) |
| Toothache | 0 (0.0) | 1 (12.5) | 0 (0.0) | 0 (0.0) | 0 (0.0) | 0 (0.0) | 0 (0.0) | 0 (0.0) | 0 (0.0) | 0 (0.0) | 0 (0.0) | 0 (0.0) | 0 (0.0) | 0 (0.0) | 1 (0.7) |
| **General disorders and administration site conditions - Total** | **6 (22.2)** | **0 (0.0)** | **0 (0.0)** | **1 (12.5)** | **0 (0.0)** | **0 (0.0)** | **0 (0.0)** | **0 (0.0)** | **1 (7.7)** | **2 (25.0)** | **4 (50.0)** | **3 (37.5)** | **3 (37.5)** | **4 (50.0)** | **24 (17.6)** |
| Application site anaesthesia | 1 (3.7) | 0 (0.0) | 0 (0.0) | 0 (0.0) | 0 (0.0) | 0 (0.0) | 0 (0.0) | 0 (0.0) | 0 (0.0) | 0 (0.0) | 0 (0.0) | 0 (0.0) | 0 (0.0) | 0 (0.0) | 1 (0.7) |
| Application site erythema | 1 (3.7) | 0 (0.0) | 0 (0.0) | 0 (0.0) | 0 (0.0) | 0 (0.0) | 0 (0.0) | 0 (0.0) | 0 (0.0) | 0 (0.0) | 0 (0.0) | 0 (0.0) | 0 (0.0) | 0 (0.0) | 1 (0.7) |
| Application site rash | 4 (14.8) | 0 (0.0) | 0 (0.0) | 1 (12.5) | 0 (0.0) | 0 (0.0) | 0 (0.0) | 0 (0.0) | 0 (0.0) | 1 (12.5) | 1 (12.5) | 0 (0.0) | 0 (0.0) | 1 (12.5) | 8 (5.9) |
| Asthenia | 0 (0.0) | 0 (0.0) | 0 (0.0) | 0 (0.0) | 0 (0.0) | 0 (0.0) | 0 (0.0) | 0 (0.0) | 0 (0.0) | 0 (0.0) | 0 (0.0) | 2 (25.0) | 0 (0.0) | 0 (0.0) | 2 (1.5) |
| Chest discomfort | 0 (0.0) | 0 (0.0) | 0 (0.0) | 0 (0.0) | 0 (0.0) | 0 (0.0) | 0 (0.0) | 0 (0.0) | 0 (0.0) | 1 (12.5) | 1 (12.5) | 0 (0.0) | 0 (0.0) | 0 (0.0) | 2 (1.5) |
| Chills | 0 (0.0) | 0 (0.0) | 0 (0.0) | 0 (0.0) | 0 (0.0) | 0 (0.0) | 0 (0.0) | 0 (0.0) | 0 (0.0) | 0 (0.0) | 0 (0.0) | 0 (0.0) | 0 (0.0) | 1 (12.5) | 1 (0.7) |
| Fatigue | 1 (3.7) | 0 (0.0) | 0 (0.0) | 0 (0.0) | 0 (0.0) | 0 (0.0) | 0 (0.0) | 0 (0.0) | 0 (0.0) | 2 (25.0) | 2 (25.0) | 2 (25.0) | 2 (25.0) | 2 (25.0) | 11 (8.1) |
| Feeling cold | 0 (0.0) | 0 (0.0) | 0 (0.0) | 0 (0.0) | 0 (0.0) | 0 (0.0) | 0 (0.0) | 0 (0.0) | 0 (0.0) | 0 (0.0) | 0 (0.0) | 0 (0.0) | 2 (25.0) | 1 (12.5) | 3 (2.2) |
| Feeling hot | 0 (0.0) | 0 (0.0) | 0 (0.0) | 0 (0.0) | 0 (0.0) | 0 (0.0) | 0 (0.0) | 0 (0.0) | 0 (0.0) | 0 (0.0) | 1 (12.5) | 0 (0.0) | 0 (0.0) | 2 (25.0) | 3 (2.2) |
| Inflammation | 1 (3.7) | 0 (0.0) | 0 (0.0) | 0 (0.0) | 0 (0.0) | 0 (0.0) | 0 (0.0) | 0 (0.0) | 0 (0.0) | 0 (0.0) | 0 (0.0) | 0 (0.0) | 0 (0.0) | 0 (0.0) | 1 (0.7) |
| Influenza like illness | 0 (0.0) | 0 (0.0) | 0 (0.0) | 0 (0.0) | 0 (0.0) | 0 (0.0) | 0 (0.0) | 0 (0.0) | 0 (0.0) | 0 (0.0) | 0 (0.0) | 0 (0.0) | 0 (0.0) | 2 (25.0) | 2 (1.5) |
| Malaise | 0 (0.0) | 0 (0.0) | 0 (0.0) | 0 (0.0) | 0 (0.0) | 0 (0.0) | 0 (0.0) | 0 (0.0) | 1 (7.7) | 0 (0.0) | 0 (0.0) | 0 (0.0) | 0 (0.0) | 0 (0.0) | 1 (0.7) |
| Pyrexia | 0 (0.0) | 0 (0.0) | 0 (0.0) | 0 (0.0) | 0 (0.0) | 0 (0.0) | 0 (0.0) | 0 (0.0) | 0 (0.0) | 0 (0.0) | 0 (0.0) | 0 (0.0) | 1 (12.5) | 1 (12.5) | 2 (1.5) |
| **Infections and infestations - Total** | **0 (0.0)** | **1 (12.5)** | **0 (0.0)** | **0 (0.0)** | **1 (12.5)** | **0 (0.0)** | **0 (0.0)** | **0 (0.0)** | **0 (0.0)** | **0 (0.0)** | **0 (0.0)** | **1 (12.5)** | **2 (25.0)** | **2 (25.0)** | **7 (5.1)** |
| Nasopharyngitis | 0 (0.0) | 1 (12.5) | 0 (0.0) | 0 (0.0) | 1 (12.5) | 0 (0.0) | 0 (0.0) | 0 (0.0) | 0 (0.0) | 0 (0.0) | 0 (0.0) | 1 (12.5) | 2 (25.0) | 1 (12.5) | 6 (4.4) |
| Oral herpes | 0 (0.0) | 0 (0.0) | 0 (0.0) | 0 (0.0) | 0 (0.0) | 0 (0.0) | 0 (0.0) | 0 (0.0) | 0 (0.0) | 0 (0.0) | 0 (0.0) | 0 (0.0) | 0 (0.0) | 1 (12.5) | 1 (0.7) |
| **Injury, poisoning and procedural complications - Total** | **0 (0.0)** | **1 (12.5)** | **1 (12.5)** | **0 (0.0)** | **0 (0.0)** | **0 (0.0)** | **0 (0.0)** | **1 (12.5)** | **0 (0.0)** | **0 (0.0)** | **0 (0.0)** | **0 (0.0)** | **0 (0.0)** | **0 (0.0)** | **3 (2.2)** |
| Contusion | 0 (0.0) | 0 (0.0) | 0 (0.0) | 0 (0.0) | 0 (0.0) | 0 (0.0) | 0 (0.0) | 1 (12.5) | 0 (0.0) | 0 (0.0) | 0 (0.0) | 0 (0.0) | 0 (0.0) | 0 (0.0) | 1 (0.7) |
| Head injury | 0 (0.0) | 1 (12.5) | 0 (0.0) | 0 (0.0) | 0 (0.0) | 0 (0.0) | 0 (0.0) | 0 (0.0) | 0 (0.0) | 0 (0.0) | 0 (0.0) | 0 (0.0) | 0 (0.0) | 0 (0.0) | 1 (0.7) |
| Limb injury | 0 (0.0) | 0 (0.0) | 1 (12.5) | 0 (0.0) | 0 (0.0) | 0 (0.0) | 0 (0.0) | 0 (0.0) | 0 (0.0) | 0 (0.0) | 0 (0.0) | 0 (0.0) | 0 (0.0) | 0 (0.0) | 1 (0.7) |
| **Investigations - Total** | **0 (0.0)** | **0 (0.0)** | **0 (0.0)** | **0 (0.0)** | **0 (0.0)** | **0 (0.0)** | **0 (0.0)** | **1 (12.5)** | **1 (7.7)** | **0 (0.0)** | **0 (0.0)** | **0 (0.0)** | **0 (0.0)** | **0 (0.0)** | **2 (1.5)** |
| Hepatic enzyme increased | 0 (0.0) | 0 (0.0) | 0 (0.0) | 0 (0.0) | 0 (0.0) | 0 (0.0) | 0 (0.0) | 1 (12.5) | 1 (7.7) | 0 (0.0) | 0 (0.0) | 0 (0.0) | 0 (0.0) | 0 (0.0) | 2 (1.5) |
| **Metabolism and nutrition disorders - Total** | **0 (0.0)** | **0 (0.0)** | **0 (0.0)** | **0 (0.0)** | **0 (0.0)** | **0 (0.0)** | **0 (0.0)** | **0 (0.0)** | **0 (0.0)** | **0 (0.0)** | **0 (0.0)** | **0 (0.0)** | **1 (12.5)** | **0 (0.0)** | **1 (0.7)** |
| Decreased appetite | 0 (0.0) | 0 (0.0) | 0 (0.0) | 0 (0.0) | 0 (0.0) | 0 (0.0) | 0 (0.0) | 0 (0.0) | 0 (0.0) | 0 (0.0) | 0 (0.0) | 0 (0.0) | 1 (12.5) | 0 (0.0) | 1 (0.7) |
| **Musculoskeletal and connective tissue disorders - Total** | **1 (3.7)** | **1 (12.5)** | **1 (12.5)** | **1 (12.5)** | **0 (0.0)** | **1 (12.5)** | **0 (0.0)** | **0 (0.0)** | **1 (7.7)** | **0 (0.0)** | **0 (0.0)** | **0 (0.0)** | **1 (12.5)** | **0 (0.0)** | **7 (5.1)** |
| Back pain | 0 (0.0) | 0 (0.0) | 0 (0.0) | 1 (12.5) | 0 (0.0) | 1 (12.5) | 0 (0.0) | 0 (0.0) | 1 (7.7) | 0 (0.0) | 0 (0.0) | 0 (0.0) | 0 (0.0) | 0 (0.0) | 3 (2.2) |
| Muscle spasms | 1 (3.7) | 0 (0.0) | 1 (12.5) | 0 (0.0) | 0 (0.0) | 0 (0.0) | 0 (0.0) | 0 (0.0) | 0 (0.0) | 0 (0.0) | 0 (0.0) | 0 (0.0) | 0 (0.0) | 0 (0.0) | 2 (1.5) |
| Muscular weakness | 0 (0.0) | 1 (12.5) | 0 (0.0) | 0 (0.0) | 0 (0.0) | 0 (0.0) | 0 (0.0) | 0 (0.0) | 0 (0.0) | 0 (0.0) | 0 (0.0) | 0 (0.0) | 0 (0.0) | 0 (0.0) | 1 (0.7) |
| Myalgia | 0 (0.0) | 0 (0.0) | 0 (0.0) | 0 (0.0) | 0 (0.0) | 0 (0.0) | 0 (0.0) | 0 (0.0) | 0 (0.0) | 0 (0.0) | 0 (0.0) | 0 (0.0) | 1 (12.5) | 0 (0.0) | 1 (0.7) |
| Pain in extremity | 0 (0.0) | 0 (0.0) | 0 (0.0) | 1 (12.5) | 0 (0.0) | 0 (0.0) | 0 (0.0) | 0 (0.0) | 0 (0.0) | 0 (0.0) | 0 (0.0) | 0 (0.0) | 0 (0.0) | 0 (0.0) | 1 (0.7) |
| **Nervous system disorders - Total** | **4 (14.8)** | **3 (37.5)** | **2 (25.0)** | **4 (50.0)** | **3 (37.5)** | **1 (12.5)** | **1 (12.5)** | **0 (0.0)** | **5 (38.5)** | **3 (37.5)** | **3 (37.5)** | **5 (62.5)** | **8 (100)** | **7 (87.5)** | **49 (36.0)** |
| Dizziness | 1 (3.7) | 0 (0.0) | 1 (12.5) | 1 (12.5) | 1 (12.5) | 1 (12.5) | 0 (0.0) | 0 (0.0) | 1 (7.7) | 1 (12.5) | 0 (0.0) | 1 (12.5) | 1 (12.5) | 3 (37.5) | 12 (8.8) |
| Dizziness postural | 1 (3.7) | 0 (0.0) | 0 (0.0) | 1 (12.5) | 0 (0.0) | 0 (0.0) | 0 (0.0) | 0 (0.0) | 0 (0.0) | 0 (0.0) | 0 (0.0) | 1 (12.5) | 1 (12.5) | 0 (0.0) | 4 (2.9) |
| Dysgeusia | 0 (0.0) | 0 (0.0) | 0 (0.0) | 0 (0.0) | 0 (0.0) | 0 (0.0) | 0 (0.0) | 0 (0.0) | 1 (7.7) | 0 (0.0) | 1 (12.5) | 0 (0.0) | 0 (0.0) | 0 (0.0) | 2 (1.5) |
| Headache | 1 (3.7) | 3 (37.5) | 0 (0.0) | 1 (12.5) | 2 (25.0) | 0 (0.0) | 0 (0.0) | 0 (0.0) | 3 (23.1) | 2 (25.0) | 2 (25.0) | 4 (50.0) | 8 (100) | 7 (87.5) | 33 (24.3) |
| Lethargy | 0 (0.0) | 0 (0.0) | 1 (12.5) | 0 (0.0) | 0 (0.0) | 0 (0.0) | 0 (0.0) | 0 (0.0) | 0 (0.0) | 0 (0.0) | 0 (0.0) | 0 (0.0) | 0 (0.0) | 0 (0.0) | 1 (0.7) |
| Paraesthesia | 0 (0.0) | 0 (0.0) | 0 (0.0) | 0 (0.0) | 0 (0.0) | 0 (0.0) | 0 (0.0) | 0 (0.0) | 0 (0.0) | 0 (0.0) | 0 (0.0) | 0 (0.0) | 0 (0.0) | 1 (12.5) | 1 (0.7) |
| Presyncope | 0 (0.0) | 0 (0.0) | 0 (0.0) | 0 (0.0) | 0 (0.0) | 0 (0.0) | 0 (0.0) | 0 (0.0) | 0 (0.0) | 1 (12.5) | 0 (0.0) | 0 (0.0) | 0 (0.0) | 0 (0.0) | 1 (0.7) |
| Restless legs syndrome | 0 (0.0) | 0 (0.0) | 0 (0.0) | 0 (0.0) | 0 (0.0) | 0 (0.0) | 0 (0.0) | 0 (0.0) | 0 (0.0) | 0 (0.0) | 0 (0.0) | 1 (12.5) | 0 (0.0) | 0 (0.0) | 1 (0.7) |
| Somnolence | 0 (0.0) | 0 (0.0) | 0 (0.0) | 0 (0.0) | 0 (0.0) | 0 (0.0) | 0 (0.0) | 0 (0.0) | 2 (15.4) | 0 (0.0) | 0 (0.0) | 0 (0.0) | 0 (0.0) | 0 (0.0) | 2 (1.5) |
| Syncope | 0 (0.0) | 0 (0.0) | 0 (0.0) | 1 (12.5) | 0 (0.0) | 0 (0.0) | 0 (0.0) | 0 (0.0) | 0 (0.0) | 0 (0.0) | 0 (0.0) | 0 (0.0) | 0 (0.0) | 0 (0.0) | 1 (0.7) |
| Tremor | 1 (3.7) | 0 (0.0) | 0 (0.0) | 0 (0.0) | 0 (0.0) | 0 (0.0) | 1 (12.5) | 0 (0.0) | 0 (0.0) | 0 (0.0) | 0 (0.0) | 0 (0.0) | 0 (0.0) | 0 (0.0) | 2 (1.5) |
| **Psychiatric disorders - Total** | **0 (0.0)** | **0 (0.0)** | **0 (0.0)** | **1 (12.5)** | **0 (0.0)** | **0 (0.0)** | **0 (0.0)** | **0 (0.0)** | **0 (0.0)** | **1 (12.5)** | **0 (0.0)** | **0 (0.0)** | **1 (12.5)** | **0 (0.0)** | **3 (2.2)** |
| Dysphoria | 0 (0.0) | 0 (0.0) | 0 (0.0) | 0 (0.0) | 0 (0.0) | 0 (0.0) | 0 (0.0) | 0 (0.0) | 0 (0.0) | 0 (0.0) | 0 (0.0) | 0 (0.0) | 1 (12.5) | 0 (0.0) | 1 (0.7) |
| Restlessness | 0 (0.0) | 0 (0.0) | 0 (0.0) | 1 (12.5) | 0 (0.0) | 0 (0.0) | 0 (0.0) | 0 (0.0) | 0 (0.0) | 1 (12.5) | 0 (0.0) | 0 (0.0) | 0 (0.0) | 0 (0.0) | 2 (1.5) |
| Tension | 0 (0.0) | 0 (0.0) | 0 (0.0) | 0 (0.0) | 0 (0.0) | 0 (0.0) | 0 (0.0) | 0 (0.0) | 0 (0.0) | 1 (12.5) | 0 (0.0) | 0 (0.0) | 0 (0.0) | 0 (0.0) | 1 (0.7) |
| **Respiratory, thoracic and mediastinal disorders - Total** | **0 (0.0)** | **0 (0.0)** | **0 (0.0)** | **0 (0.0)** | **2 (25.0)** | **0 (0.0)** | **0 (0.0)** | **0 (0.0)** | **1 (7.7)** | **1 (12.5)** | **3 (37.5)** | **0 (0.0)** | **1 (12.5)** | **3 (37.5)** | **11 (8.1)** |
| Cough | 0 (0.0) | 0 (0.0) | 0 (0.0) | 0 (0.0) | 1 (12.5) | 0 (0.0) | 0 (0.0) | 0 (0.0) | 0 (0.0) | 0 (0.0) | 0 (0.0) | 0 (0.0) | 0 (0.0) | 1 (12.5) | 2 (1.5) |
| Dry throat | 0 (0.0) | 0 (0.0) | 0 (0.0) | 0 (0.0) | 0 (0.0) | 0 (0.0) | 0 (0.0) | 0 (0.0) | 0 (0.0) | 0 (0.0) | 0 (0.0) | 0 (0.0) | 1 (12.5) | 0 (0.0) | 1 (0.7) |
| Dyspnoea | 0 (0.0) | 0 (0.0) | 0 (0.0) | 0 (0.0) | 0 (0.0) | 0 (0.0) | 0 (0.0) | 0 (0.0) | 1 (7.7) | 0 (0.0) | 3 (37.5) | 0 (0.0) | 0 (0.0) | 1 (12.5) | 5 (3.7) |
| Nasal discomfort | 0 (0.0) | 0 (0.0) | 0 (0.0) | 0 (0.0) | 0 (0.0) | 0 (0.0) | 0 (0.0) | 0 (0.0) | 0 (0.0) | 1 (12.5) | 0 (0.0) | 0 (0.0) | 0 (0.0) | 0 (0.0) | 1 (0.7) |
| Oropharyngeal pain | 0 (0.0) | 0 (0.0) | 0 (0.0) | 0 (0.0) | 1 (12.5) | 0 (0.0) | 0 (0.0) | 0 (0.0) | 0 (0.0) | 0 (0.0) | 0 (0.0) | 0 (0.0) | 0 (0.0) | 0 (0.0) | 1 (0.7) |
| Rhinorrhoea | 0 (0.0) | 0 (0.0) | 0 (0.0) | 0 (0.0) | 1 (12.5) | 0 (0.0) | 0 (0.0) | 0 (0.0) | 0 (0.0) | 0 (0.0) | 0 (0.0) | 0 (0.0) | 0 (0.0) | 0 (0.0) | 1 (0.7) |
| Sleep apnoea syndrome | 0 (0.0) | 0 (0.0) | 0 (0.0) | 0 (0.0) | 0 (0.0) | 0 (0.0) | 0 (0.0) | 0 (0.0) | 0 (0.0) | 0 (0.0) | 0 (0.0) | 0 (0.0) | 0 (0.0) | 1 (12.5) | 1 (0.7) |
| **Skin and subcutaneous tissue disorders - Total** | **2 (7.4)** | **0 (0.0)** | **1 (12.5)** | **0 (0.0)** | **0 (0.0)** | **0 (0.0)** | **1 (12.5)** | **0 (0.0)** | **4 (30.8)** | **1 (12.5)** | **0 (0.0)** | **0 (0.0)** | **0 (0.0)** | **0 (0.0)** | **9 (6.6)** |
| Dermatitis contact | 0 (0.0) | 0 (0.0) | 1 (12.5) | 0 (0.0) | 0 (0.0) | 0 (0.0) | 0 (0.0) | 0 (0.0) | 3 (23.1) | 0 (0.0) | 0 (0.0) | 0 (0.0) | 0 (0.0) | 0 (0.0) | 4 (2.9) |
| Hyperhidrosis | 1 (3.7) | 0 (0.0) | 0 (0.0) | 0 (0.0) | 0 (0.0) | 0 (0.0) | 1 (12.5) | 0 (0.0) | 0 (0.0) | 0 (0.0) | 0 (0.0) | 0 (0.0) | 0 (0.0) | 0 (0.0) | 2 (1.5) |
| Rash | 1 (3.7) | 0 (0.0) | 0 (0.0) | 0 (0.0) | 0 (0.0) | 0 (0.0) | 0 (0.0) | 0 (0.0) | 1 (7.7) | 0 (0.0) | 0 (0.0) | 0 (0.0) | 0 (0.0) | 0 (0.0) | 2 (1.5) |
| Skin irritation | 1 (3.7) | 0 (0.0) | 0 (0.0) | 0 (0.0) | 0 (0.0) | 0 (0.0) | 0 (0.0) | 0 (0.0) | 0 (0.0) | 1 (12.5) | 0 (0.0) | 0 (0.0) | 0 (0.0) | 0 (0.0) | 2 (1.5) |
| **Vascular disorders - Total** | **0 (0.0)** | **0 (0.0)** | **0 (0.0)** | **0 (0.0)** | **0 (0.0)** | **0 (0.0)** | **0 (0.0)** | **0 (0.0)** | **3 (23.1)** | **0 (0.0)** | **0 (0.0)** | **0 (0.0)** | **0 (0.0)** | **0 (0.0)** | **3 (2.2)** |
| Haematoma | 0 (0.0) | 0 (0.0) | 0 (0.0) | 0 (0.0) | 0 (0.0) | 0 (0.0) | 0 (0.0) | 0 (0.0) | 1 (7.7) | 0 (0.0) | 0 (0.0) | 0 (0.0) | 0 (0.0) | 0 (0.0) | 1 (0.7) |
| Orthostatic hypotension | 0 (0.0) | 0 (0.0) | 0 (0.0) | 0 (0.0) | 0 (0.0) | 0 (0.0) | 0 (0.0) | 0 (0.0) | 2 (15.4) | 0 (0.0) | 0 (0.0) | 0 (0.0) | 0 (0.0) | 0 (0.0) | 2 (1.5) |
